# Senotherapeutic role of pemafibrate through autophagy/mitophagy regulation in chronic obstructive pulmonary disease

**DOI:** 10.64898/2026.08.31.26361865

**Authors:** Sachi Matsubayashi-Hosokawa, Saburo Ito, Yusuke Hosaka, Masahiro Yoshida, Tsukasa Kadota, Mitsuo Hashimoto, Satoki Hatano, Tomoya Maruyama, Shota Fujimoto, Saiko Nishioka, Shun Inukai, Yu Fujita, Shunsuke Minagawa, Hiromichi Hara, Takeo Nakada, Katsutoshi Nakayama, Takashi Ohtsuka, Kazuyoshi Kuwano, Jun Araya

**Author notes:** Correspondence and requests for reprints should be addressed to: Saburo Ito, M.D., Ph.D., The Jikei University School of Medicine, 3-25-8, Nishi-Shimbashi, Minato-ku, Tokyo, 105-8461, Japan, Jun Araya, M.D., Ph.D., The Jikei University School of Medicine, 3-25-8, Nishi-Shimbashi, Minato-ku, Tokyo, 105-8461, Japan. These authors contributed equally to this work (Sachi Matsubayashi-Hosokawa, Saburo Ito).

## Abstract

Inadequate autophagy promotes smoking-induced cellular senescence involved in chronic obstructive pulmonary disease (COPD) pathogenesis. Transcription factor EB (TFEB) is a master regulator of the autophagy-lysosome axis. For the first time, we investigated the therapeutic potential of pemafibrate, a putative TFEB inducer. COPD lung epithelial cells showed reduced TFEB expression. Pemafibrate enhanced autophagy/mitophagy flux and restored lysosomal acidification observed during cigarette smoke (CS) extract exposure in human bronchial epithelial cells, resulting in reduced cellular senescence. TFEB knockdown demonstrated involvement of pemafibrate-induced TFEB in these effects. Pemafibrate induced TFEB expression, mitigated alveolar enlargement and airflow obstruction, and attenuated the CS-induced increase in static lung compliance in a long-term CS-exposed mouse model. It reduced the CS exposure-induced cellular senescence, possibly through autophagy/mitophagy, as suggested by bulk RNA sequencing of mouse lungs. A retrospective cohort study showed that patients given pemafibrate displayed attenuated FEV_1.0_ decline compared with those given bezafibrate or fenofibrate. In conclusion, pemafibrate is a promising therapeutic agent for COPD, potentially exerting its effects through the regulation of the TFEB–autophagy/mitophagy–lysosome axis.

## Introduction

Chronic obstructive pulmonary disease (COPD) is mainly caused by long-term cigarette smoke (CS) exposure and is characterized by progressive airflow limitation(1, 2). Advanced age is one of the most important risk factors for COPD development, and recent research advances have implicated accelerated senescence in a variety of cell types, including epithelial cells, fibroblasts, and lymphocytes, during COPD pathogenesis(1). Cellular senescence is a hallmark of aging, and accumulation of senescent cells plays a pivotal role in the aging process of organ dysfunction, through impaired cell regeneration and aberrant cytokine secretion in the senescence-associated secretory phenotype(3, 4).

Autophagy is an evolutionarily conserved degradation pathway that targets cytoplasmic components, including macromolecules and organelles(5). Owing to the highly selective nature of cellular clearance, autophagy is linked to the maintenance of cell and tissue homeostasis by preventing defective organelle accumulation and misfolded protein aggregates(5). Autophagic activity declines with age and has been widely implicated in aging and longevity(5–7). Dysregulation of autophagy, including impaired lysosomal fusion and/or degradation, contributes to the mechanisms underlying cellular senescence progression(5, 8). Senotherapies targeting cellular senescence, including senomorphics and senolytics, can delay or attenuate aging-related diseases, and autophagy has been recognized as an essential mechanism for senotherapeutic action(3, 9–11).

Insufficient autophagy has been implicated in accelerated cellular senescence in COPD lungs(12). Damaged mitochondria in COPD lungs were observed by electron microscopic evaluation(13). We previously demonstrated that phosphatase and tensin homolog deleted from chromosome 10 (PTEN)-induced putative kinase 1 (PINK1)-Parkin RBR E3 ubiquitin protein ligase (PRKN) pathway-mediated mitophagy plays a key regulatory role in mitochondrial reactive oxygen species (ROS) production and cellular senescence induced by CS extract (CSE) in human bronchial epithelial cells (HBECs)(14). Reduced PRKN expression levels in COPD lungs and an exaggerated COPD phenotype in a CS-exposed PRKN knockout mouse model further indicated that insufficient mitophagy is part of the pathogenic sequence of COPD progression, and it acts by modulating cellular senescence(14–16).

The functional integrity of lysosomes is critical for preserving the autophagy machinery. Lysosomal damage, represented by lysosomal membrane permeabilization and/or rupture, can be caused by a variety of cellular stressors, including silica, oxidative stress, and lysosomotropic agents(17–19). Lysosomal damage stimulates a specific autophagic response known as lysophagy, which eliminates damaged lysosomes and maintains lysosomal integrity(20, 21). We have reported lysosomal damage in terms of lysosomal membrane permeabilization (with concomitant lysosomal dysfunction in epithelial cells) in association with cellular senescence in COPD lungs, which can be attributed to impaired lysophagy(22). Accordingly, insufficient autophagy/mitophagy due to dysregulation of the autophagy-lysosome axis possibly plays a pivotal role in accelerating cellular senescence in COPD pathogenesis.

Transcription factor EB (TFEB) is a master regulator of the autophagy-lysosome axis and belongs to the microphthalmia/transcription factor E (MiT/TFE) family of basic helix-loop-helix leucine zipper transcription factors(23). TFEB is involved in various cellular processes, including energy homeostasis, metabolism autophagy-lysosomal biogenesis, and cellular senescence (23, 24). The lack of MiT/TFE transcription factors results in impaired degradation of damaged mitochondria, suggesting functional crosstalk between TFEB and mitophagy for regulating mitochondrial integrity during stress conditions(25). TFEB dysregulation has been implicated in the pathogenesis of various diseases, including lysosomal storage disorders, neurodegenerative diseases, and hepatic steatosis(23). TFEB activation-mediated restoration of the autophagy-lysosome axis has been postulated to be a promising therapeutic approach for these disorders(23). Sequestered TFEB in cytoplasmic aggresome bodies has been observed in lung tissues, and TFEB malfunction may be partly responsible for insufficient autophagy in COPD pathogenesis(26). Intriguingly, the peroxisome proliferator-activated receptor alpha (PPARα) agonist gemfibrozil has been shown to salvage the perinuclear accumulation of TFEB aggregates caused by CSE stimulation and suppress alveolar epithelial cell death(26). In addition, a recent database analysis study showed that another PPARα agonist, fenofibrate, reduced the incidence of chronic diseases associated with aging and increased life expectancy, in part through functional alteration of bone marrow senescence(27). Accordingly, we hypothesized that pemafibrate, a selective peroxisome proliferator-activated receptor α modulator (SPPARMα) which has more specific pharmacological effect than gemfibrozil and fenofibrate, may regulate TFEB, and thereby serve as a promising senotherapeutic approach for patients with COPD who have a dysregulated autophagy-lysosome axis.

In the present study, we examined TFEB expression levels in COPD samples and evaluated the regulatory role of TFEB in CS-induced autophagy/mitophagy activation and cellular senescence. Furthermore, the therapeutic potential of pemafibrate was examined in terms of TFEB and mitophagy modulation in cell cultures and CS-exposed mouse models *in vivo*. We also attempted to elucidate the potential preventive role of pemafibrate against the decline in pulmonary function in a retrospective cohort study.

## Results

### TFEB is involved in autophagy and cellular senescence regulation during CSE exposure

To clarify the role of TFEB in COPD pathogenesis, we first examined changes in protein levels and nuclear translocation upon CSE exposure. Treatment of HBECs with CSE for 24 h caused a significant increase in TFEB protein levels (Fig. 1A). However, the induction of TFEB expression by CSE decreased to the same level or lower than that before stimulation at 48 h, suggesting that the TFEB induction by CSE stimulation is transient. This result is consistent with our previous finding that autophagy induction by CSE is transient and insufficient autophagy is associated with accumulation of damaged intracellular organelles such as mitochondria, resulting in increased ROS levels, DNA damage, and cellular senescence during CSE exposure(14). The down-shift of TFEB observed upon CSE stimulation may reflect its activation through phosphorylation, as previously reported (Fig. 1A) (17). Nuclear translocation of TFEB activation was also detected following CSE exposure, in a dose-dependent manner, in enhanced green fluorescent protein (EGFP)-TFEB-expressing BEAS-2B cells, which is a representative bronchial epithelial cell line (Fig. 1B). TFEB is a master regulator of the autophagy-lysosome axis, suggesting that CSE-activated TFEB regulates autophagy-and lysosome-related genes. We used a small interfering RNA (siRNA) against TFEB for TFEB knockdown and carried out a quantitative polymerase chain reaction (qPCR) to confirm that it efficiently reduced the TFEB mRNA levels. TFEB knockdown clearly suppressed autophagy-associated *BECN1* and *VPS18*, as well as lysosome-associated *ATP6V1A* and *LAMP1*, suggesting the involvement of TFEB in autophagic flux during CS exposure (Fig. 1C). Consistent with previous findings(12), CSE induced autophagy activation, as shown by increased EGFP-microtubule-associated protein 1A/1B light chain 3 (LC3) dot formation and microtubule-associated proteins 1A/1B light chain 3B (LC3B) conversion from LC3B-I to LC3B-II; this activation was suppressed by TFEB knockdown (Fig. 1D,E). We previously reported the pivotal role of autophagy in regulating CSE-induced cellular senescence(12). TFEB knockdown significantly enhanced the CSE-induced cellular senescence, as shown by means of p21/cyclin-dependent kinase inhibitor 1A (CDKN1A) expression and phosphohistone H2A.X (Ser139) staining, suggesting the participation of TFEB-mediated autophagy flux in regulating CSE-induced HBEC senescence (Fig. 1F,G).

**Figure 1.**
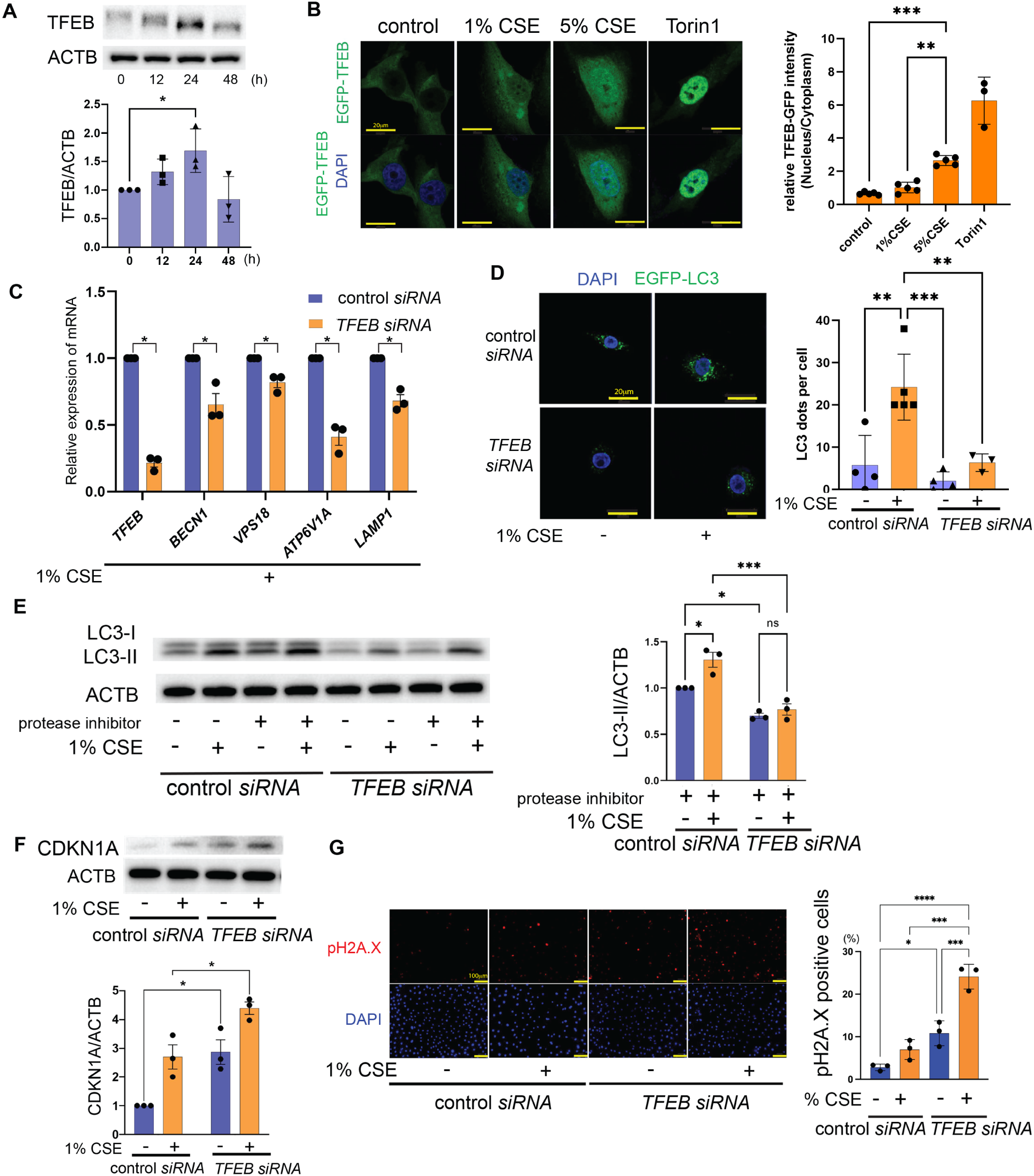
TFEB regulates autophagy and cellular senescence following CSE exposure. HBECs or BEAS-2B cells were transfected using siRNA at 24h before CSE treatment. (**A**) Western blot using anti-TFEB and -ACTB antibodies, on lysates of HBECs subjected to control or 1% CSE treatment for the indicated times. The lower panel shows relative expression obtained upon densitometry (n = 3). (**B**) Fluorescence microscopy images of DAPI and EGFP-TFEB in BEAS-2B cells, after 24 h of treatment with CSE (1%–5%). Torin 1 (250 nM, 6h) was used as a positive control. DAPI (blue) and TFEB (green) are shown. The right panel shows the relative nuclear-to-cytoplasmic EGFP-TFEB intensity ratio (n = 5; Torin 1, n = 3). Scale bar: 20 µm. (**C**) Real-time PCR analysis of TFEB and autophagy- and lysosome-related gene expression in HBECs treated with 1% CSE for 12 hours. Expression was normalized to ACTB and is shown relative to the control siRNA group. (n = 3). (**D**) Confocal laser scanning microscopic images of DAPI and EGFP-LC3 in BEAS-2B cells, after 24 h of treatment with CSE (1%). EGFP-LC3 BEAS-2B cells were transfected with either control or *TFEB siRNA*. The right panel shows the number of LC3 dots per cell (n = 3-5). Scale bar: 20 µm. (**E**) Western blot of HBEC cell lysates using anti-LC3B and -ACTB antibodies. HBECs were transfected with either control or *TFEB siRNA*, and the cell lysates were collected after 24 h of treatment with CSE (1%). Protease inhibitor (E64d, 10 mg/ml, pepstatin A, 10 mg/ml) were added 6 h before sample collection. The right panels show the relative expression determined by densitometry (n = 3). (**F**) Western blot of HBEC cell lysates using anti-CDKN1A (p21) and -ACTB antibodies. HBECs were transfected with either control or *TFEB siRNA*, and the cell lysates were collected after 24 h of treatment with CSE (1%). The right panels show the relative expression determined by densitometry (n = 3). (**G**) Fluorescence microscopic images of DAPI and phosphorylated histone H2A.X (pH2A.X) in HBECs, after 24 h of treatment with CSE (1%). HBECs were transfected with either control or *TFEB siRNA*. Scale bar: 100 µm. Shown in the right panel is the percentage of pH2A.X-positive cells (*n*=3). Data are presented as mean ± SEM with individual data points. \**p*<0.05, \*\**p*<0.01, \*\*\**p*<0.001, and \*\*\*\**p*<0.0001. Student’s *t*-test was used for comparisons between two groups, whereas one-way or two-way ANOVA followed by Bonferroni’s multiple comparisons test was used for comparisons among multiple groups, as appropriate, unless otherwise indicated.

### TFEB expression is reduced in COPD lungs

Although we observed the involvement of TFEB in CS-induced cellular senescence in HBECs, TFEB expression levels in COPD lungs remain obscure. GEO2R analysis of the microarray dataset GSE106986, which compares gene transcription in the whole lungs of 14 patients with COPD and 5 non-smokers, showed a significant reduction in TFEB expression among patients with COPD (Fig. 2A). Another microarray dataset, GSE994, which shows gene transcription in airway epithelial cells isolated from 23 non-smokers, 18 former smokers, and 34 current smokers, showed a significant reduction in TFEB expression in the current smokers (Fig. 2B). We also examined TFEB expression in small airway epithelial cells using immunohistochemistry. The percentage of positive nuclear staining for TFEB was calculated in non-smokers (33 airways from six patients), non-COPD smokers (26 airways from five patients), and patients with COPD (43 airways from seven patients), which revealed a significant reduction in the percentage of TFEB-positive nuclei in the patients with COPD (Fig. 2C). Our recent comprehensive single-cell transcriptome evaluation revealed the importance of the phenotypic alteration of epithelial cells, represented by inflammatory alveolar type 2 cells, in COPD pathogenesis(28). Using this dataset, we examined *TFEB* and *autophagy-related gene* (*ATG*) expression in lung epithelial cells. Compared with lung epithelial cells from non-smokers (three patients) and non-COPD smokers (four patients), epithelial cells from patients with COPD (nine patients) showed significant reduction in *TFEB*, *ATG7,* and *ATG10* levels and a trend of reduction in *ATG5* levels (Fig. 2D). We also checked the expression of *TFEB* and *ATGs* in whole lung tissue of the same dataset. Although significant differences were not observed in both *TFEB* and *ATGs* due to their low expression levels, the expression of *ATG5*, *ATG7*, and *ATG10* tended to be consistently lower in the condition of COPD in all lung cells (Fig. S1). A significant reduction in the expression of *TFEB*, *ATG7*, and *ATG10* in COPD epithelial cells was further confirmed by quantitative RT-PCR in HBECs isolated from patients (Fig. 2E). This result suggests that the reduced *TFEB* and *ATGs* expression observed in the COPD lung epithelium is causally linked to insufficient autophagy associated with accelerated epithelial cell senescence during COPD pathogenesis.

**Figure 2.**
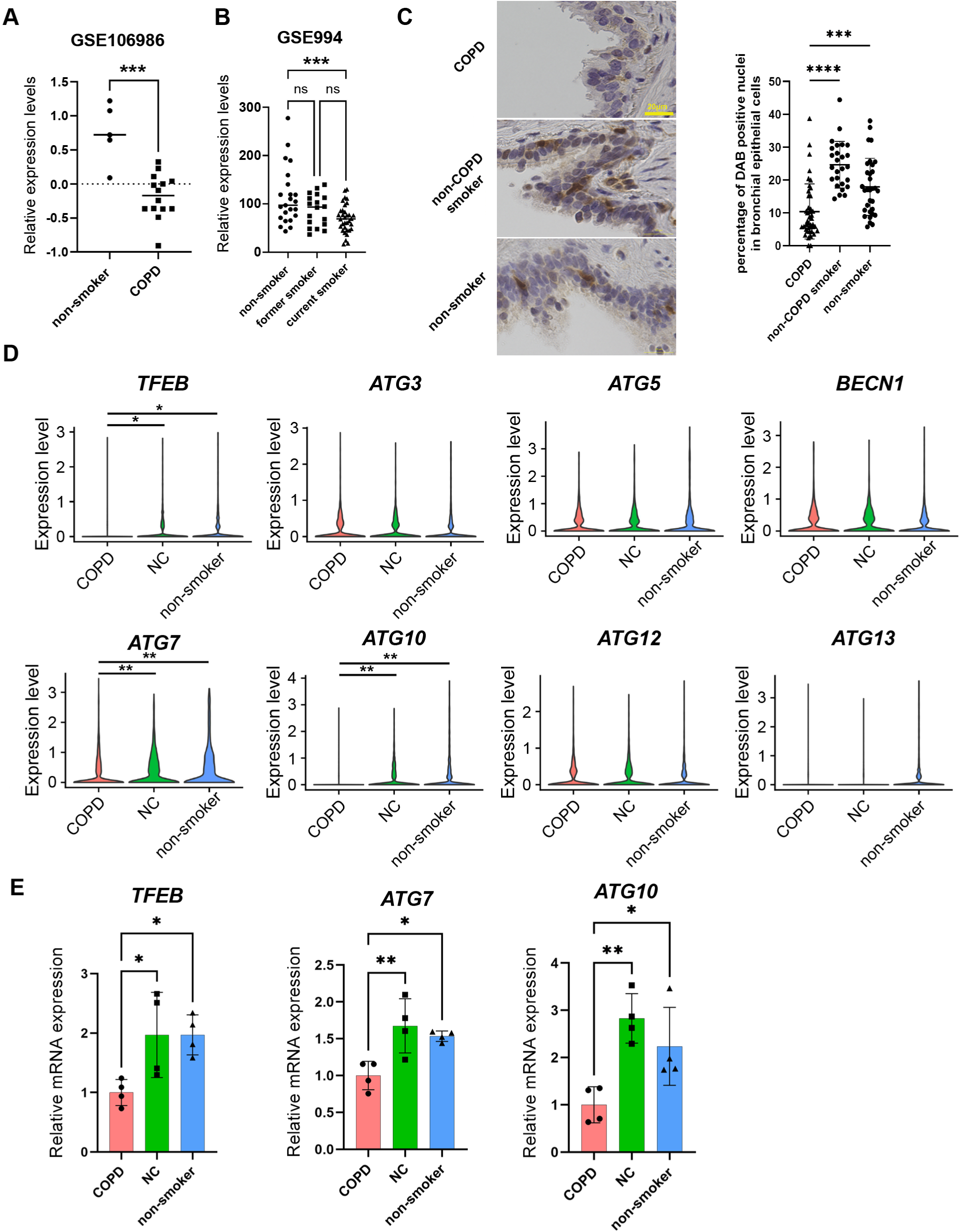
*TFEB* and other autophagy-related gene expression in COPD and non-COPD lungs. (A and B). GEO2R analysis of microarray dataset. (**A**) *TFEB* expression in the whole lungs of patients with COPD and non-smokers, using the GSE106986 microarray dataset. (**B**) *TFEB* expression in the whole lungs of current smokers, former smokers, and non-smokers, using the GSE994 microarray dataset. (**C**) Immunohistochemical staining for TFEB in the airways of surgically resected lungs. The right panel shows the percentage of TFEB-positive cell nuclei in the airway epithelial cells. Patients with COPD: 7 samples, 43 airways; non-COPD smokers: 5 samples, 26 airways; non-smokers: 6 samples, 33 airways. Scale bar: 20 µm. (**D**) *TFEB* and autophagy-related gene expression in lung epithelial cells from patients with COPD, non-COPD smokers, and nonsmokers in a previously published single-cell transcriptomic dataset(28). Lung epithelial cells were analyzed based on the cell type annotations provided in the original study. (**E**) Real-time PCR analysis of TFEB, ATG7, ATG10 expression in surgically resected lung tissues. Relative mRNA expression was calculated after normalization to ACTB and is shown relative to the indicated control group. (n = 4 per group). \**p*<0.05, \*\**p*<0.01, \*\*\**p*<0.001, and \*\*\*\**p*<0.0001. For comparisons among three groups, one-way ANOVA followed by Bonferroni post-hoc test was used. For two-group comparisons, Student’s *t*-test was used. NC, non-COPD smokers.

### Pemafibrate induces TFEB expression and enhances autophagy flux, concomitantly restoring lysosomal acidity and mitochondrial function in response to CSE exposure

A PPARα agonist is generally used for the treatment of dyslipidemia(29) and PPARα-mediated TFEB induction has been demonstrated upon treatment(30). Pemafibrate is now used for the treatment of dyslipidemia because of its high efficacy and safety compared to other fibrates, due to its high selectivity and pharmacological effect for PPARα(31). We took notice of the potential efficacy of pemafibrate in the COPD pathogenesis of accelerated cellular senescence through the activation of TFEB and autophagy. First, we examined the effect of pemafibrate on TFEB expression in airway epithelial cells. Pemafibrate induced TFEB expression in a dose-dependent manner, and significant induction was detected upon treatment of BEAS-2B cells with 10 nM pemafibrate and that of HBECs with 100 nM pemafibrate (Fig. 3A). Additionally, simultaneous CSE and pemafibrate treatment induced higher degree of TFEB nuclear translocation than that observed in CSE alone (Fig. S2A). In contrast to CSE, pemafibrate alone failed to induce a down-shift of TFEB or evident nuclear translocation, indicating that pemafibrate-mediated upregulation of TFEB is insufficient to trigger its activation under cell culture conditions lacking CSE exposure (Fig. 3A and S2A). We then demonstrated the involvement of PPARα in pemafibrate-mediated TFEB expression by means of PPARα knockdown experiments (Fig. S2B). Pemafibrate induced autophagy activation in the setting of CSE exposure, as shown in terms of increased EGFP-LC3B dot formation (Fig. 3B). We also examined the effects of pemafibrate on mitophagy. Co-localization of mitochondria stained with translocase of outer mitochondrial membrane 20 (TOMM20) and EGFP-LC3B puncta was used to assess mitophagy(14). Pemafibrate significantly enhanced mitophagy, particularly under conditions of CSE exposure (Fig. 3C). Increased co-localization of TOMM20 with LAMP1 further supported pemafibrate-mediated mitophagy (Fig. S2C). TFEB is a master regulator of the autophagy–lysosomal axis. To evaluate not only autophagic flux but also lysosomal acidity, which can be impaired in COPD pathogenesis(22), BEAS-2B cells expressing the mRFP-GFP tandem fluorescent-tagged LC3 (tfLC3) construct were used. CSE treatment alone increased both yellow and green puncta, with only modest elevations in the flux index (red/yellow puncta ratio). In contrast, pemafibrate markedly increased the flux index together with an increased red/green fluorescence ratio (autolysosomal acidity), suggesting that pemafibrate enhanced autophagic flux and restored lysosomal acidity during CSE exposure (Fig. 3D). Pemafibrate-mediated lysosomal acidification was further confirmed using LysoSensor Yellow/Blue DND-160 and LysoTracker™ Red staining (Fig. 3E and Fig. S2D). Next, we sought to determine whether pemafibrate-induced mitophagy contributed to the restoration of mitochondrial dysfunction. CSE-induced mitochondrial damage, indicated by mitochondrial ROS production, was significantly attenuated by pemafibrate treatment, as shown by MitoSOX™ Red staining (Fig. 3F). This effect was abrogated by PPARα knockdown (Fig. S2E). To assess mitochondrial turnover, BEAS-2B cells expressing mito-Dendra2 were locally photo-converted from green to red fluorescence. The red fluorescence intensity declined after 3 h of CSE exposure, and this decline was further enhanced in the presence of pemafibrate, indicating that pemafibrate promoted mitochondrial degradation through mitophagy (Fig. 3G). Mitochondrial respiration was evaluated by measuring the oxygen consumption rate (OCR) using a Seahorse XF Flux Analyzer (Agilent Technologies). Exposure to CSE resulted in a significant decrease in basal and maximal respiration, consistent with mitochondrial dysfunction. This impairment was restored by pemafibrate treatment (Fig. 3H). Mitochondrial functional restoration was further demonstrated by MitoTracker™ Red CMXRos staining, which showed that pemafibrate mitigated CSE-induced depolarization of the mitochondrial membrane potential (Fig. S3A). No significant changes in antioxidant pathways, including GPx and GSH, were demonstrated by pemafibrate treatment (Fig. S3B). These results suggest that mitochondrial quality control mechanisms, particularly mitophagy rather than antioxidant pathways, are primarily responsible for maintaining mitochondrial functional integrity and regulating mitochondrial ROS production during pemafibrate treatment. Pemafibrate also mitigated the CSE exposure-induced cellular senescence in HBECs, as demonstrated by means of positive senescence-associated β-galactosidase (SA-β-Gal) staining intensity (Fig. 3I).

**Figure 3.**
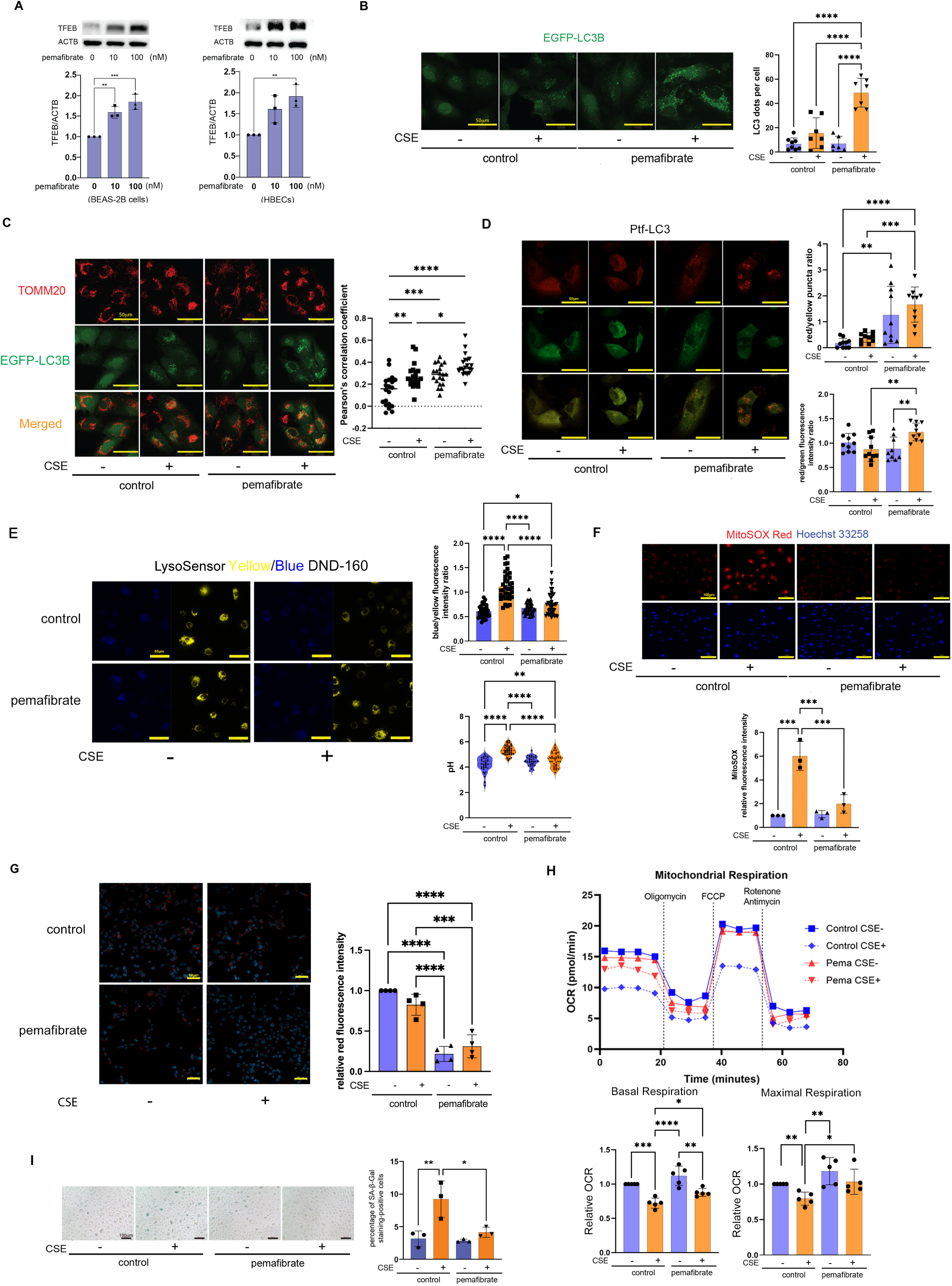
Pemafibrate mitigates CS-induced cellular senescence via autophagy/mitophagy induction. (**A**) Western blot of lysates from BEAS-2B cells (left panel) and HBECs (right panel), using anti-TFEB and - ACTB antibodies. Protein samples were collected after 24 h of treatment with pemafibrate (0–100 nM). The lower panels show the relative expression obtained upon densitometric analysis of the western blot (*n*=3). (**B**) Confocal laser scanning microscopic images of EGFP-LC3 in BEAS-2B cells, after 24 h of treatment with CSE (2%). Pemafibrate (100 nM) was added 24 h before CSE treatment. The right panel shows number of LC3 dots per cell. LC3 puncta per cell were quantified in 7–8 randomly selected cells per condition. Scale bar: 50 µm. (**C**) Co-localization analysis of TOMM20 and EGFP-LC3B in BEAS-2B cells, after 24 h of treatment with CSE (2%), using confocal laser scanning microscopy. Pemafibrate (100 nM) was added 24 h before CSE treatment. The right panel shows the quantification of co-localization based on Pearson’s correlation coefficient which were calculated within randomly selected cell ROIs. Scale bar: 50 µm. (**D**) Confocal laser scanning microscopic images of Ptf-LC3 in BEAS-2B cells, after 24 h of treatment with CSE (2%). Pemafibrate (100 nM) was added 24 h before CSE treatment. The upper right panel shows the red/yellow puncta ratio, and the lower right panel shows the red/green fluorescence intensity ratio (10 cells per condition). Quantification was performed in randomly selected cells. (**E**) Lysosomal pH was assessed by ratiometric live-cell imaging using LysoSensor Yellow/Blue DND-160 (2 μM, 5 min). Representative confocal images show yellow (510–560 nm) and blue (430–470 nm) channel fluorescence under each condition. The blue/yellow fluorescence intensity ratio (upper right) and calculated lysosomal pH (lower right) are shown. Data are presented as mean ± SEM with individual data points. (**F**) Hoechst 33258 and MitoSOX™ Red fluorescence staining of HBECs, after 24 h of treatment with CSE (2%). Pemafibrate (100 nM) was added 24 h before CSE treatment. The right panel shows the relative fluorescence intensity of MitoSOX. Fluorescence intensity was normalized to the control group (n = 3 independent experiment). Scale bar: 100 µm. (**G**) Confocal laser scanning microscopic images of mitoDendra2 in BEAS-2B cells, after 3 h of treatment with CSE (2%). Pemafibrate (100 nM) was added 24 h before CSE treatment. Red fluorescence intensity of photoconverted mitoDendra2 was quantified within cell ROIs using ImageJ. Fluorescence intensity was normalized to the control group, which was set to 1, and expressed as relative fluorescence intensity. (n =3 independent experiments). (**H**) Mitochondrial respiration in HBECs exposed to 2% CSE for 60 minutes, with or without 100 nM pemafibrate pretreatment for 24 hours. Oligomycin (1.5 µM), FCCP (2 µM), and rotenone/antimycin A (0.5 µM) were sequentially injected as indicated. OCR traces are presented as the averages of three technical replicates. The lower left panel shows the quantification of basal respiration, expressed as relative OCR (control = 1.0), and the lower right panel shows maximal respiration (n = 5, independent experiments). (**I**) Photomicrographs of SA-β-Gal staining of HBECs. HBECs were treated with 1% CSE for 24 h. Pemafibrate (100 nM) was added 24 h before CSE treatment. The lower panel shows the percentage of SA-β-Gal-positive cells (n = 3). Data are presented as mean ± SEM with individual data points. \**p*<0.05, \*\**p*<0.01, and \*\*\**p*<0.001. Student’s *t*-test was used for comparisons between two groups, whereas one-way or two-way ANOVA followed by Bonferroni’s multiple comparisons test was used for comparisons among multiple groups, as appropriate, unless otherwise indicated.

To confirm the participation of pemafibrate-induced TFEB in maintaining the integrity of the mitophagy-lysosome axis, we performed TFEB knockdown experiments. TFEB knockdown suppressed the pemafibrate-induced activation of mitophagy during CSE exposure (Fig. 4A). Pemafibrate-mediated suppression of mitochondrial ROS production and restoration of lysosomal acidification were also abrogated by TFEB knockdown (Fig. 4B, C). In addition, TFEB knockdown mitigated the anti-senescence effects of pemafibrate (Fig. 4D).

**Figure 4.**
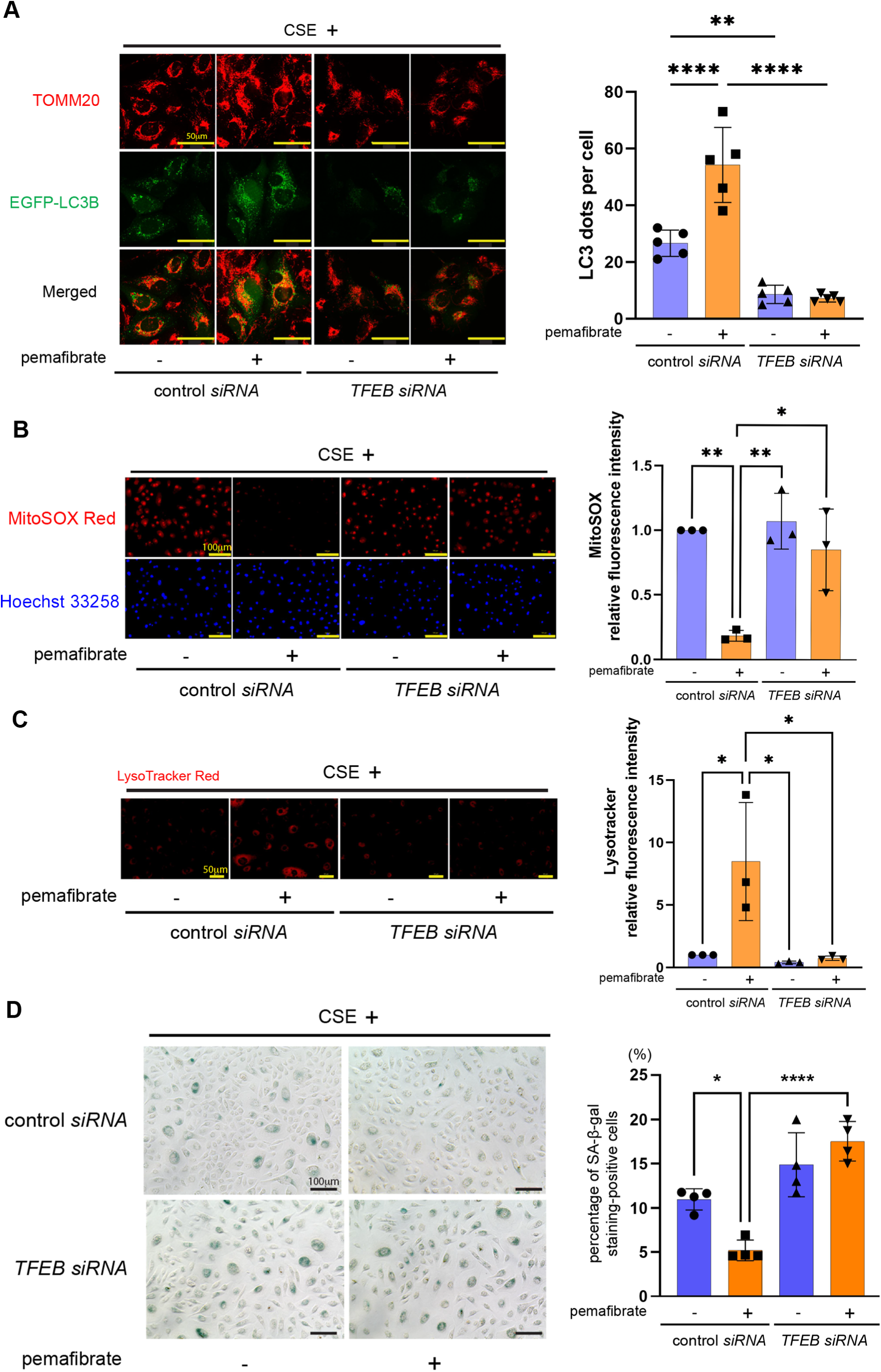
TFEB knockdown attenuates pemafibrate-mediated mitophagy activation in CS-treated bronchial epithelial cells. BEAS-2B cells or HBECs were first transfected with control or *TFEB siRNA*, and then 24 h later, treated with pemafibrate (100 nM), for 24 h. Finally, the cells received treatment with CSE (2%) for 24 h. (**A**) Co-localization analysis of TOMM20 and EGFP-LC3B in BEAS-2B cells, using confocal laser scanning microscopy. The number of EGFP-LC3B dots per cell was quantified and shown on the right (n = 5). Scale bar: 50 µm. (**B**) MitoSOX™ Red and Hoechst 33258 fluorescence staining of the HBECs. The right panel shows the relative MitoSOX fluorescence intensity, normalized to the control group, is shown on the right (n = 3 independent experiments). Scale bar: 100 μm. (**C**) LysoTracker™ Red fluorescence staining of the HBECs. The right panel shows the relative fluorescence intensity of lysotracker. Fluorescence intensity was normalized to the control siRNA/CSE(−) group. Scale bar: 50 µm. (**D**) SA-β-Gal staining of the HBECs. Shown in the right panel is the percentage of SA-β-Gal-positive cells (*n*=4). Scale bar: 100 µm. Data are presented as mean ± SEM with individual data points. \**p*<0.05, \*\**p*<0.01, and \*\*\*\**p*<0.0001, as analyzed using analysis of variance and Bonferroni *post-hoc* test.

### Pemafibrate attenuates emphysematous changes in a long-term CS exposure mouse model

To elucidate the potential efficacy of pemafibrate as a novel COPD treatment, a long-term (6 months) CS exposure mouse model was used, and pemafibrate was administered to it through food. CS exposure induced emphysematous changes in the mean linear intercept (Fig. 5A). Decreased forced expiratory volume in 0.1 second to inspiratory capacity ratio (FEV₀.₁/IC), reflecting airway obstruction, and increased static lung compliance (Cst) indicated the establishment of a COPD phenotype following long-term CS exposure in our mouse model (Fig. 5B). These COPD-related phenotypic changes were significantly mitigated by pemafibrate treatment (Fig. 5A, B). Although small airways are thought to contribute substantially to airway obstruction in COPD pathogenesis, alterations in the small airways observed under our experimental conditions were relatively modest (Fig. S4A). Pemafibrate treatment significantly attenuated the elevated total and macrophage cell counts in the bronchoalveolar lavage fluid of the CS-exposed group, as compared with those observed in the control group (Fig. 5C). It also significantly reduced the CS exposure-induced cellular senescence, which was assessed in terms of enhanced expression levels of p21/CDKN1A and p16/cyclin-dependent kinase inhibitor 2A (CDKN2A), and positive SPiDER-β-Gal staining (Fig. 5D,E). In CS-exposed mice, bronchial epithelial cells exhibited morphological alterations characterized by shortened mitochondrial length and disrupted cristae, which were significantly alleviated by pemafibrate treatment (Fig. S4B). Accordingly, pemafibrate may attenuate the COPD phenotype by preventing accelerated cellular senescence that results from impaired mitophagy.

**Figure 5.**
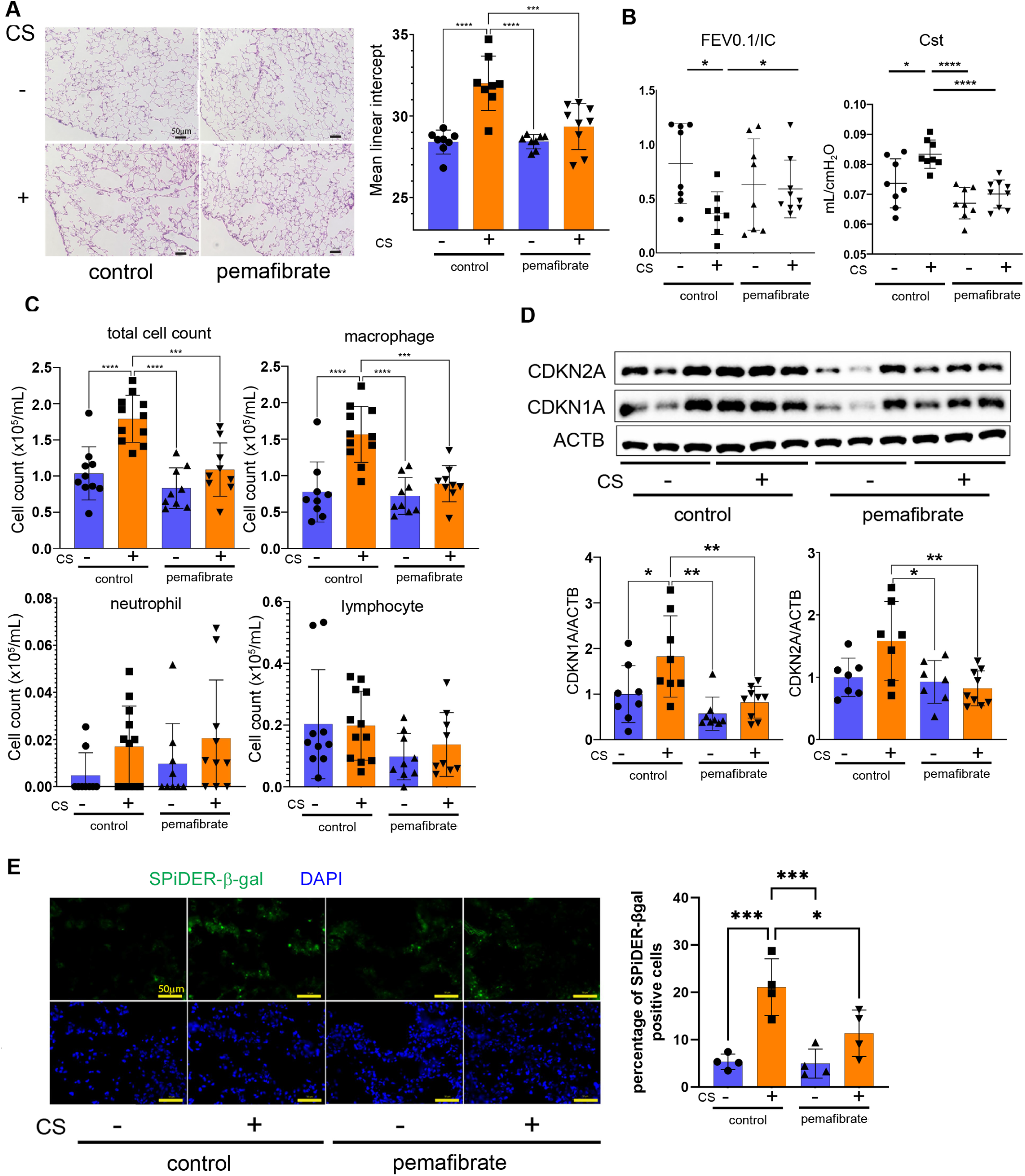
Pemafibrate attenuates cigarette smoke-induced emphysematous changes and cellular senescence *in vivo*. Mice were exposed to room air or cigarette smoke for six months with or without pemafibrate administration. (**A**) Representative lung sections and mean linear intercept measurements. Five randomly selected fields were analyzed for each mouse, and the average value for each mouse was used as one biological replicate. Mean linear intercept measurements are shown on the right. *n*=8 (mice) in the control diet:room air (CR) group; *n*=8 in the control diet: CS exposure (CC) group; *n*=8 in the pemafibrate-mixed diet:room air (PR) group; and *n*=9 in the pemafibrate-mixed diet:CS exposure (PC) group. Scale bar: 50 µm. (**B**) Respiratory function in mice. The left and right panels show the FEV_0.1_/IC ratio and static lung compliance (Cst), respectively. Parameters are shown for each individual mouse. *n*=8 in the CR group, *n*=8 in the CC group, *n*=8 in the PR group, and *n*=9 in the PC group. (**C**) Cell counts of total cells, macrophages, neutrophils, and lymphocytes in the bronchoalveolar lavage fluid from mice. *n*=10 in the CR group, *n*=11 in the CC group, *n*=9 in the PR group, and *n*=9 in the PC group. (**D**) Western blot showing the expression levels of CDKN2A, CDKN1A, and ACTB in the homogenized mice lungs. Relative expression determined by densitometry is shown in the lower panel. *n*=8 in the CR group, *n*=8 in the CC group, *n*=8 in the PR group, and *n*=9 in the PC group. (**E**) SPiDER-β-Gal and DAPI staining of the mice lungs. Scale bar: 50 µm. The right panel shows the percentage of SPiDER-β-Gal-positive cells. Multiple randomly selected fields were analyzed for each mouse, and the average value for each mouse was used as one biological replicate. n = 4 mice per group. Data are presented as mean ± SEM with individual data points. \**p*<0.05, \*\**p*<0.01, \*\*\**p*<0.001, and \*\*\*\**p*<0.0001, as analyzed using analysis of variance and Bonferroni *post-hoc* test.

### Pemafibrate induces the expression of TFEB and its target genes in a long-term CS exposure mouse model

Immunohistochemical evaluation of lungs from CS-exposed mice treated with pemafibrate demonstrated a significant increase in nuclear TFEB staining in airway epithelial cells (Fig. 6A). In whole-lung samples, pemafibrate treatment tended to increase TFEB expression at both the mRNA and protein levels, whereas a significant induction was observed only when combined with CS exposure (Fig. 6B, C). To enable a comprehensive evaluation, bulk RNA sequencing (RNA-seq) was performed on whole-lung samples from mice. Consistent with the protein data, RNA-seq analysis revealed a trend toward increased expression of both TFEB following pemafibrate treatment (Fig. 6D). To evaluate transcriptional activity of TFEB induced by pemafibrate, expression levels of coordinated lysosomal expression and regulation (CLEAR) network was assessed(32). Clearly increased CLEAR network gene expression was demonstrated in heatmap (Fig. 6E). The transcriptional activity of TFEB was indirectly assessed using the mean Z-scores of CLEAR network genes, suggesting that pemafibrate significantly enhances TFEB transcriptional activity regardless of CS exposure (Fig. 6F).

**Figure 6.**
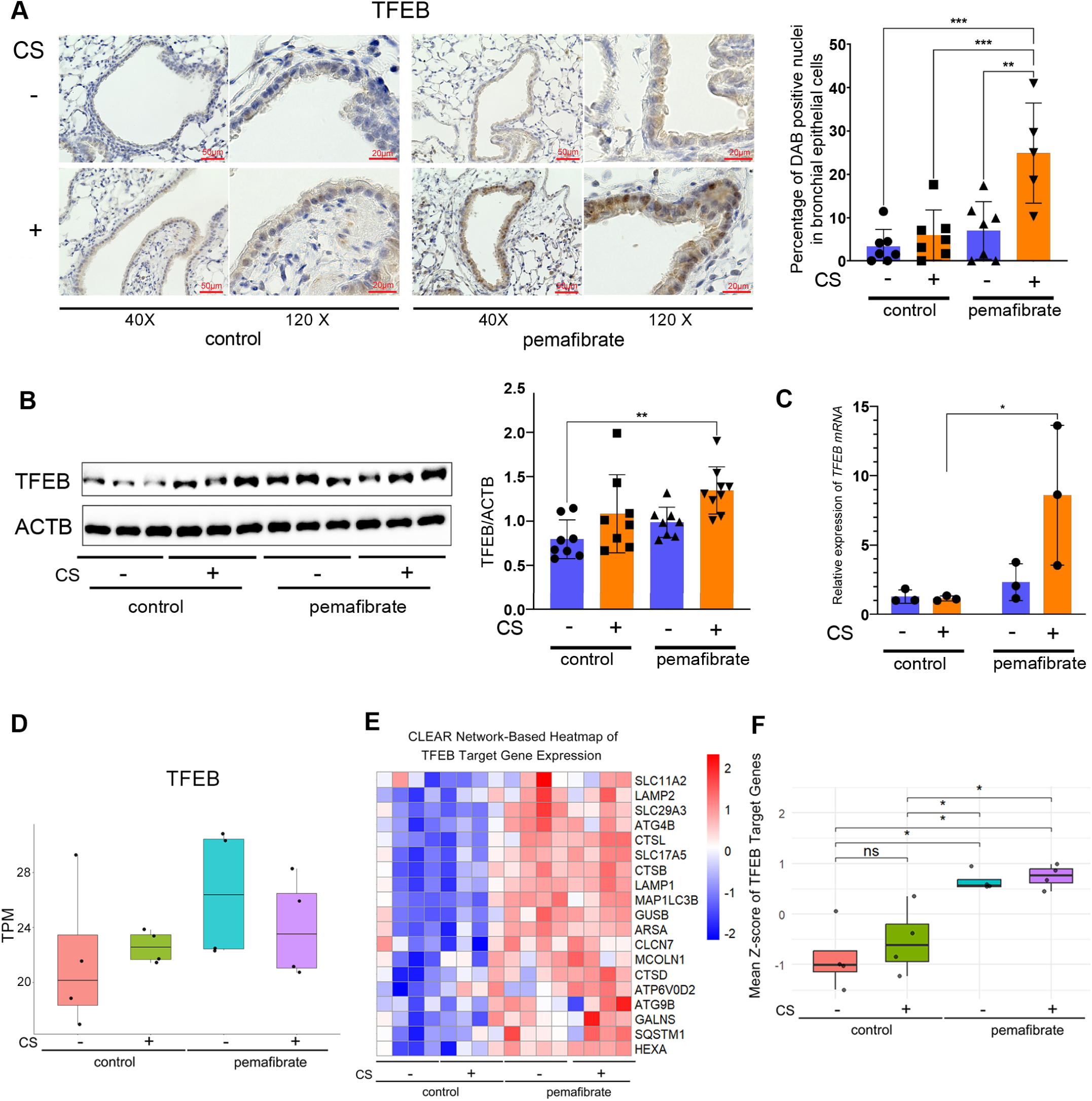
Pemafibrate induces TFEB activation in the lungs of CS-exposed mice. (**A**) Immunohistochemical staining for TFEB in the mice lung airways. The right panel shows the percentage of TFEB-positive cell nuclei in the airway epithelial cells. Multiple airways were analyzed for each mouse, and the average value for each mouse was used as one biological replicate. *n*=7 in the control diet:room air group; *n*=7 in the control diet: CS exposure group; *n*=7 in the pemafibrate-mixed diet:room air group; and *n*=5 in the pemafibrate-mixed diet:CS exposure group. Scale bars: 50 µm (40×) and 20 µm (120×). (**B**) Representative western blots of the expression levels of TFEB and ACTB in the mice lung homogenates. The right panel shows the average (±SEM) relative expression obtained upon densitometric analysis of the western blot, *n*=8 in the control diet:room air group; *n*=8 in the control diet: CS exposure group; *n*=8 in the pemafibrate-mixed diet:room air group; and *n*=9 in the pemafibrate-mixed diet:CS exposure group. (**C**) Real-time PCR was performed using primers for *TFEB* and *ACTB* (as control). Relative mRNA expression was calculated after normalization to ACTB. (*n*=3 per group). (**D**) Box plots show transcript levels (TPM) of TFEB in four experimental conditions: control cells without CS exposure, control cells with CS exposure, pemafibrate-treated cells without CS exposure, and pemafibrate-treated cells with CS exposure. RNA-seq was performed using whole-lung samples. (n = 4 mice per group). (**E**) CLEAR Network-based heatmap of RNA-seq results showing differences in the expression of TFEB target genes (*n*=4 per group). (**F**) Mean Z-score of TFEB target genes in RNA-seq. \**p*<0.05, \*\**p*<0.01, and \*\*\**p*<0.001, as assessed using analysis of variance and Bonferroni *post-hoc* test. NES, normalized enrichment score.

### Pemafibrate induces the expression of autophagy-related genes in a long-term CS exposure mouse model

A comprehensive comparison of gene expression changes corresponding to each step of autophagy revealed that pemafibrate significantly induced the expression of autophagy-related genes, as demonstrated by both heatmap analysis and comparisons of overall and process-specific Z-scores (Fig. 7A, B, C). To further clarify the biological significance of pemafibrate treatment, we utilized gene set enrichment analysis (GSEA) between control/CS and pemafibrate/CS groups. GSEA demonstrated that autophagy was significantly enriched and that mitophagy tended to be enriched in the lungs of pemafibrate groups (Fig. 7D). In addition, increased expression of the representative autophagy- and mitophagy-related proteins ATG5 and PINK1 was demonstrated by immunohistochemistry in airway epithelial cells and by western blotting in whole lung homogenates, respectively (Fig. S5). The causal link between TFEB expression and autophagy activation during pemafibrate treatment was further supported by a positive correlation between the expression levels of TFEB (mean TPM) and the autophagy activity score (mean Z-score of autophagy-related gene expression) (Fig. 7E). These findings support the involvement of TFEB in pemafibrate-induced autophagy and mitophagy, which may be further enhanced in the setting of CS exposure.

**Figure 7.**
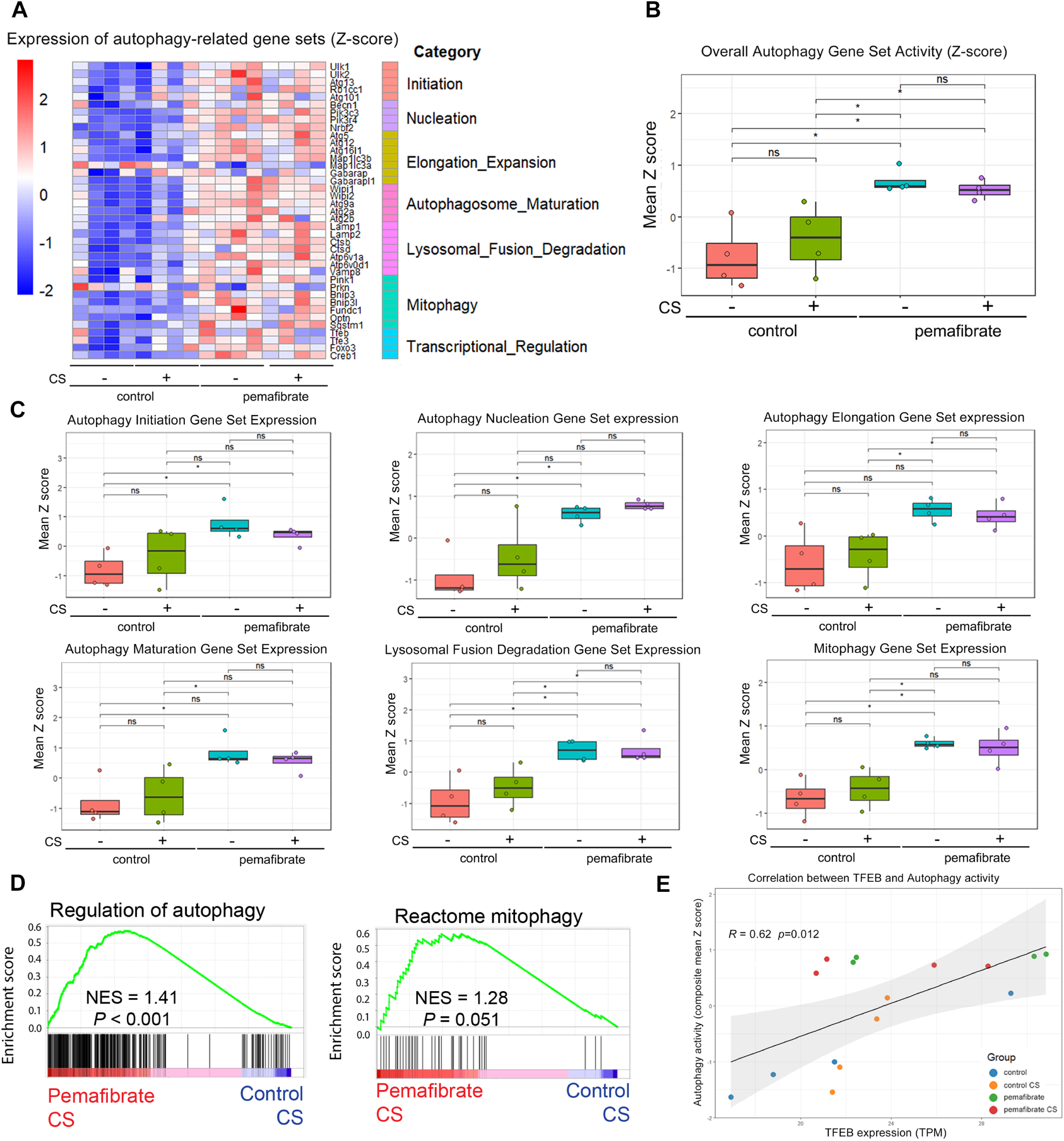
Pemafibrate activates autophagy, especially mitophagy, through the induction of TFEB in the lungs of CS-exposed mice. (**A**) Heatmap of RNA-seq results showing differences in the expression of autophagy-related genes (*n*=4 per group). (**B**) Overall autophagy gene set activity in the RNA-seq dataset was assessed using Z-scores. (**C**) Expression of gene sets corresponding to each stage of autophagy, and mitophagy in the RNA-seq dataset was evaluated using Z-scores. (**D**) Enrichment analysis of regulation of autophagy gene and reactome mitophagy gene in RNA-seq data between pemafibrate-mixed diet:CS exposure group and control diet:CS exposure group. (**E**) Correlation between TFEB and autophagy activity. Autophagy activity was calculated as the composite mean Z-score of autophagy-related genes, and TFEB expression is shown as TPM. The solid line and shaded area indicate the fitted linear regression line and 95% confidence interval. Pearson’s correlation \**p*<0.05, as assessed using analysis of variance and Bonferroni *post-hoc* test.

### Pemafibrate effectively suppresses age-related decline in pulmonary function

The forced expiratory volume in one second (FEV₁.₀) is a representative parameter of airway obstruction and is known to decline naturally with age; this decline is further accelerated during COPD development. Based on our in vitro and in vivo findings, we hypothesize that pemafibrate, a drug prescribed for the treatment of dyslipidemia, may help prevent FEV₁.₀ decline in clinical settings. To evaluate this potential preventive effect, we conducted a retrospective cohort study and analyzed clinical data obtained from patient records of individuals who were prescribed fibrates. Of 4,447 patients screened, 265 had at least 2 spirometry measurements. After excluding 19 patients with less than 4 weeks of fibrate treatment and 66 with an annualized FEV1.0 change exceeding ±180 mL/year, 180 patients were included: 26 in the pemafibrate group, 101 in the bezafibrate group, and 53 in the fenofibrate group (Figure 8A).

**Figure 8.**
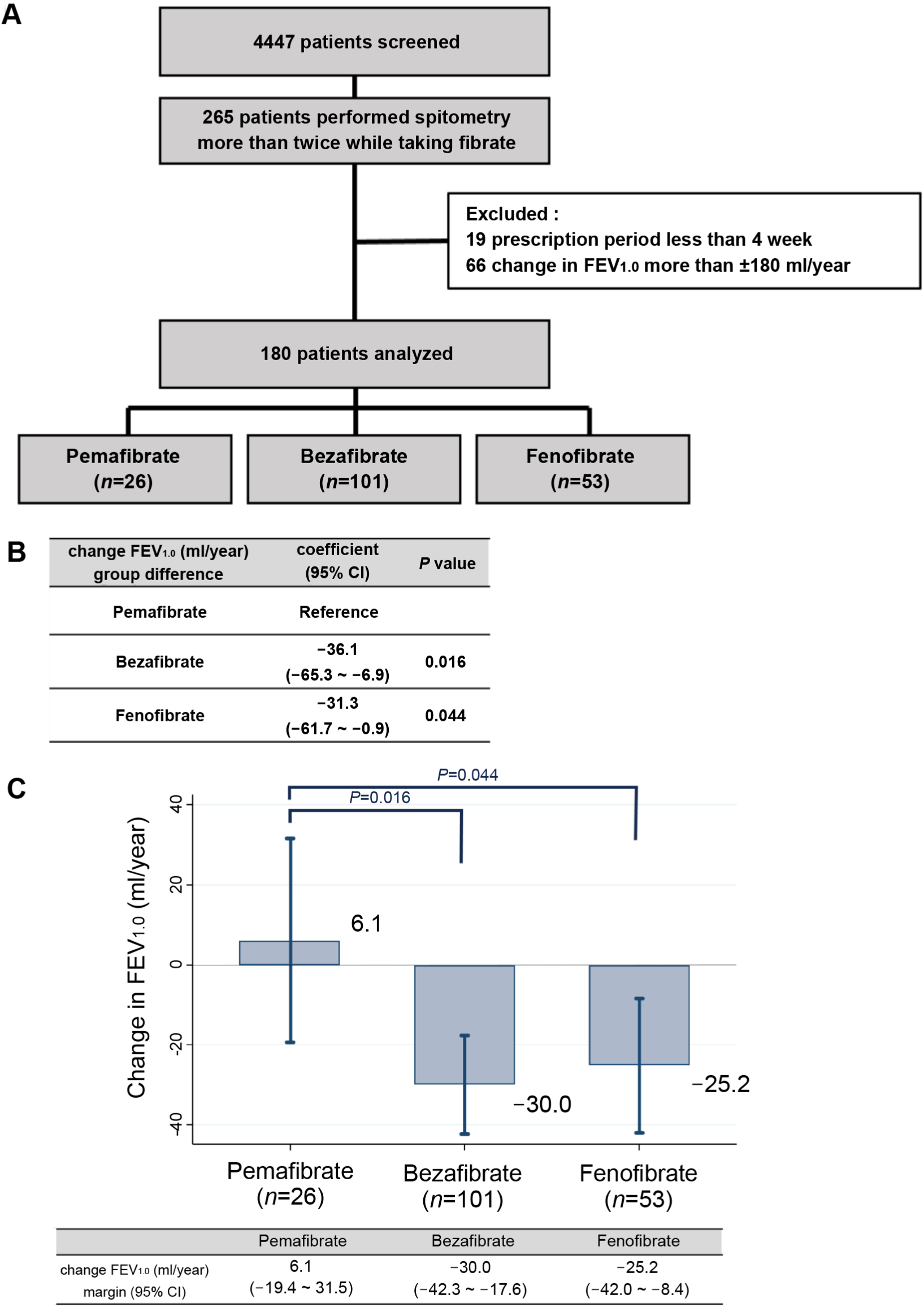
Retrospective cohort study comparing the effects of fibrate on pulmonary function. (**A**) Study design. Forty-four hundred and forty-seven individuals taking fibrates were screened, out of which 180 were included in the final analysis. (**B**) Adjusted group differences in annualized FEV1.0 change compared with the pemafibrate group. Pemafibrate was used as the reference group. Coefficients, 95% confidence intervals, and p values were estimated using analysis of covariance (ANCOVA). Age, sex, BMI, smoking index, baseline FEV_1.0_, presence of comorbidities (COPD, asthma, hypertension, diabetes, and coronary artery disease), use of statins and metformin, and use of inhalation therapy were adjusted as covariates. (**C**) Adjusted annualized change in FEV1.0 in each fibrate group. Statistical value of change in FEV_1.0_ ml per year (box) and 95% CI range (capped line).

Patient baseline characteristics are shown in Table 1 and Table S1. Baseline pulmonary functions are shown in Tables S2. The pemafibrate group included more individuals with diabetes and had a higher frequency of statin and metformin use. However, other characteristics—such as age, body mass index (BMI), respiratory comorbidities, and baseline FEV₁.₀—did not differ significantly among the pemafibrate, bezafibrate, and fenofibrate groups. In the adjusted analysis of covariance (ANCOVA) model, with the pemafibrate group used as the reference, the bezafibrate group showed a significantly greater annualized decline in FEV1.0 than the pemafibrate group (coefficient, −36.1 mL/year; 95% CI, −65.4 to −6.9; p = 0.016). Similarly, the fenofibrate group showed a significantly greater annualized decline in FEV1.0 than the pemafibrate group (coefficient, −31.3 mL/year; 95% CI, −61.7 to −0.9; p = 0.044) (Fig. 8B). A margin plot showing the adjusted annualized change in FEV1.0 for each group is shown in Fig. 8C.

**Table 1.** Baseline characteristics of each fibrate group (*n*=180).

| | | Pemafibrate<br>($n=26$) | | Bezafibrate<br>($n=101$) | | Fenofibrate<br>($n=53$) | | Total<br>($n=180$) | | *P* value |
| --- | --- | --- | --- | --- | --- | --- | --- | --- | --- | --- |
|  |  | *n* | % | *n* | % | *n* | % | *n* | % |  |
| sex | female | 7 | 26.9 | 36 | 35.6 | 19 | 35.9 | 62 | 34.4 | 0.68 |
| age (year), mean (SD) |  | 63.4 (10.4) |  | 64.9 (13.0) |  | 63.1 (12.7) |  | 64.1 (12.5) |  | 0.55 |
| observation period (year), median (IQR) |  | 1.92 (1.02 - 3.41) |  | 2.36 (1.12 - 3.99) |  | 1.91 (0.99 - 3.29) |  | 2.19 (1.04 - 3.96) |  | 0.24 |
| BMI, mean (SD) |  | 27.7 (5.2) |  | 25.2 (4.0) |  | 26.5 (5.8) |  | 26.0 (4.8) |  | 0.069 |
| baseline FEV1.0 mL, mean (SD) |  | 2268 (659.2) |  | 2073 (657.6) |  | 2213 (762.0) |  | 2143 (691.0) |  | 0.30 |
| respiratory<br>comorbidities | COPD | 9 | 34.6 | 21 | 20.8 | 10 | 18.9 | 40 | 22.2 | 0.25 |
|  | bronchial asthma | 6 | 23.1 | 24 | 23.8 | 12 | 22.6 | 42 | 23.3 | 0.99 |
| inhalation therapy |  | 7 | 26.9 | 22 | 21.8 | 13 | 24.5 | 42 | 23.3 | 0.83 |
| other metabolic<br>disease drugs | statin | 10 | 38.5 | 7 | 6.9 | 5 | 9.4 | 22 | 12.2 | 0.000 |
|  | metformin | 8 | 30.8 | 7 | 6.9 | 9 | 17.0 | 24 | 13.3 | 0.004 |
| other<br>comorbidities | hypertension | 19 | 73.1 | 63 | 62.4 | 38 | 71.7 | 120 | 66.7 | 0.38 |
|  | diabetes | 16 | 61.5 | 27 | 26.7 | 29 | 54.7 | 72 | 40.0 | 0.000 |
|  | coronary artery disease | 4 | 15.4 | 8 | 7.9 | 3 | 5.7 | 15 | 8.3 | 0.33 |
| smoking pack-year, median (IQR) |  | 33.9 (7.5 - 41) |  | 20 (0 - 40) |  | 20 (0 - 40) |  | 22 (0 - 40) |  | 0.26 |

## Discussion

The current study demonstrates that TFEB plays a regulatory role in autophagy- and lysosome-related gene expression, autophagy/mitophagy activation, and cellular senescence during CSE exposure. TFEB expression was significantly reduced in COPD lungs, particularly in epithelial cells. Pemafibrate induced TFEB expression, resulting in a reduction of CS-induced mitochondrial ROS production, accompanied by improved mitochondrial function, restoration of lysosomal acidity, and prevention of cellular senescence. Pemafibrate mitigated the development of a CS-induced COPD phenotype in association with activating the TFEB–autophagy/mitophagy axis. Furthermore, a retrospective cohort study indicated that pemafibrate prevents age-related FEV₁.₀ decline. Overall, pemafibrate administration may represent a promising senotherapeutic option for patients with COPD who exhibit accelerated FEV₁.₀ decline through activation of the TFEB–autophagy/mitophagy-lysosome axis.

Autophagic activity declines with age through several mechanisms, including age-dependent reduction in autophagy-related genes, impaired delivery of cargo to lysosomes, and lysosomal proteolytic dysfunction(5). Compromised autophagy has been proposed as the cardinal feature of organismal aging and upregulation of autophagy promotes longevity in animals(5). Increasing basal autophagy flux by disrupting BECN1, the BCL-2 complex, has been shown to extend the lifespan and healthspan of mice(6). ATG5 overexpression enhances autophagy, resulting in an extended lifespan in mice, accompanied by tolerance to oxidative stress(33), suggesting the induction of canonical ATG expression as a therapeutic approach for aging-associated disorders. Among *Atg* genes, age-dependent reduction has been demonstrated in *ATG5*, *ATG7*, and *BECN1* in human samples(34). We previously reported that autophagy is insufficient in the lungs of patients with COPD, which can be causally associated with CS-induced cellular senescence(12). Our single-cell transcriptome evaluation demonstrated a significant reduction in *ATG7* and *ATG10* levels in lung epithelial cells from patients with COPD, further supporting the notion that impaired autophagy is involved in COPD pathogenesis. Since deterioration of lysosomes can also be responsible for decreased autophagy in COPD lungs(22), it is likely that ATG induction may not be sufficient for the restoration of autophagic activity and focused on TFEB as the master transcriptional regulator of autophagy and lysosomal system. Based on the GEO2R analysis of the microarray datasets, immunohistochemical evaluation, and single-cell transcriptome evaluation, a significant reduction in TFEB expression levels was observed in COPD. In contrast to COPD lungs, our in vitro and in vivo models showed increased TFEB expression in response to CS exposure. We speculate several possibilities to explain this discrepancy. In our in vitro experiments, we used primary HBECs from non-COPD donors and immortalized BEAS-2B cells subjected to short-term CSE exposure. These cell types or exposure conditions may not have been sufficient to recapitulate the epigenetic alterations that underlie TFEB reduction in COPD. Similarly, in our in vivo model, CS exposure was initiated at 8 weeks of age and continued until 32 weeks, indicating that the chronological age of the animals may not have been appropriate for evaluating TFEB dysregulation associated with chronic COPD progression. Given the reduction of TFEB observed in COPD lungs, we reason that therapeutic activation of TFEB may represent an effective strategy for mitigating COPD pathogenesis.

Based on the previous reports showing PPARα agonist-induced transactivation of TFEB and senotherapeutic property of TFEB modulation in COPD mouse model(26, 35), we considered that pemafibrate may be a promising senomorphic agent by promoting TFEB expression. PPARα mainly regulates metabolic pathways, including activation of fatty acid β-oxidation and apolipoprotein expression, its anti-inflammatory and -fibrotic mechanisms have also been demonstrated under experimental conditions(36–38). Pemafibrate has high selectivity for PPARα and activates PPARα at low doses compared to other fibrates(31). Compared with gemfibrozil, which has been reported to ameliorate COPD pathogenesis via TFEB induction^22^, pemafibrate exhibited a stronger anti-senescence effect by suppressing CSE-induced CDKN1A upregulation in HBECs (Fig. S6). Both our in vitro and in vivo models demonstrated that pemafibrate induced TFEB expression in lung epithelial cells. Unlike CSE-induced TFEB expression, which exhibited a down-shift indicative of activation, pemafibrate treatment did not produce such a shift, implying that pemafibrate alone is insufficient to activate TFEB under short-term in vitro conditions. Intriguingly, in our RNA sequencing analysis of mouse models subjected to long-term pemafibrate administration, the activation of CLEAR network genes mediated by TFEB was significantly enhanced in the pemafibrate-treated groups, irrespective of CS exposure (Fig. 6E, F). This suggests that long-term pemafibrate treatment may be sufficient not only to induce TFEB expression but also to promote its activation. TFEB and TFE3 both belong to the MiT/TFE transcription factor family and are known to function in a complementary manner. Our RNA-seq analysis showed a trend toward increased expression of both TFEB and TFE3 following pemafibrate administration (data not shown), suggesting a potential cooperative role of these transcription factors in pemafibrate-induced activation of CLEAR network gene expression(39). This possibility warrants further investigation in future studies. The qPCR and bulk RNA-seq data demonstrated comparable trends in TFEB expression, although the results were not entirely concordant. This discrepancy is most likely due to differences in the samples selected for each assay (Fig.6C, D). The activation of TFEB via dephosphorylation is regulated by several mechanisms, including the mechanistic target of rapamycin complex 1 (mTORC1) and AMP-activated protein kinase (AMPK), both of which respond to various signals such as nutrient availability(40). Therefore, it is plausible that long-term pemafibrate-induced alterations in nutrient status could modulate mTORC1 and AMPK activity, thereby contributing to the activation of TFEB, as observed in our mouse models.

Recent advances have revealed the highly selective nature of autophagy and the existence of a variety of selective autophagy subtypes(5). Among these, mitochondria-selective mitophagy for the maintenance of mitochondrial integrity has been widely implicated in the regulation of the aging process, including cellular senescence(41). We have previously reported that CSE induces mitophagy(14), and in the current study, we found that pemafibrate further enhanced mitophagy, potentially involving TFEB activation, in response to CSE exposure. It has been reported that TFEB displays transcriptional activity in a PINK1- and PRKN-dependent manner during mitophagy(25). Our comprehensive analysis of autophagy pathway by RNA-seq demonstrated significant elevation of autophagy-related gene expression, including mitophagy gene set by pemafibrate (Fig. 7A). Since mitophagy activation generally requires prior mitochondrial damage, it is likely that smoking-related stimuli enhance mitophagy activity. Indeed, in HBECs, mitophagy was more prominently induced by CSE exposure than by pemafibrate treatment alone, as evidenced by the co-localization of TOMM20 with LC3 and mito-Dendra2 turnover (Fig. 3C, G). Mitochondrial function was improved, as demonstrated by reduced mitochondrial ROS production and OCR restoration by flux analyzer measurements in HBECs (Fig. 3F, H). In addition, RNA-seq analysis of mouse lungs revealed an increasing trend in the expression of mitochondrial electron transport chain (ETC) genes following pemafibrate treatment, with significant upregulation observed in complexes II and V (Fig. S7), further supporting the restoration of mitochondrial functional integrity. Although mitochondrial biogenesis is an essential component of mitochondrial integrity, no significant upregulation of peroxisome proliferator-activated receptor gamma coactivator (PGC)-related genes was observed in RNA-seq analysis (data not shown), suggesting limited evidence for enhanced mitochondrial biogenesis. Intriguingly, RNA-seq analysis of mouse lungs revealed a trend toward increased expression of NRF2 in the pemafibrate-treated group (data not shown), raising the possibility that mechanisms other than TFEB—such as NRF2-mediated autophagy activation or enhanced antioxidant capacity—may be involved. However, no significant changes were observed in antioxidant pathways, including glutathione and glutathione peroxidase levels (Fig. S3B). Excessive mitophagy may also be involved in the pathogenesis of COPD *via* necroptosis, which can be attributed to increased PINK1 expression(42). Although our RNA sequencing data and immunohistochemistry showed a trend toward increased PINK1 expression in the pemafibrate-treated groups, pemafibrate treatment did not enhance cell death shown by terminal deoxynucleotidyl transferase dUTP nick end labeling (TUNEL) staining, even after prolonged CS exposure in our mouse models (Fig. S8). We speculate that pemafibrate-mediated mitophagy is primarily activated during CS exposure, and that this process may optimally prevent CS-induced cellular senescence and mitigate the development of the COPD phenotype without triggering excessive regulated cell death.

Lysosomal dysfunction has also been implicated in both cellular senescence and the pathogenesis of COPD(22). Indeed, CSE exposure induced lysosomal dysfunction characterized by reduced lysosomal acidity, as demonstrated by both tfLC3-expressing cell experiments and LysoTracker™ Red staining, which were restored by pemafibrate treatment (Fig. 3D, S2D). TFEB knockdown attenuated this effect (Fig. 4C), supporting the involvement of TFEB in pemafibrate-mediated restoration of lysosomal function. Together with the RNA-seq findings (Figure 7A), these results suggest that restoration of the autophagy–lysosome axis contributes to the senotherapeutic effects of pemafibrate in COPD.

This retrospective cohort study indicated that pemafibrate may prevent the decline in FEV_1.0_ during aging. COPD is characterized by an accelerated decline in FEV_1.0_, which results at least partly from enhanced cellular senescence. Because the number of individuals with COPD in this cohort was insufficient for statistical evaluation, we focused on the naturally occurring decline in FEV1.0 irrespective of smoking status. Although several studies have reported beneficial effects of metabolic drugs such as metformin and statins on COPD exacerbation and the decline in FEV_1.0_(43–45), few have demonstrated the favorable effects of fibrates(46). ANCOVA adjusted for clinically relevant covariates showed that pemafibrate was associated with a slower annualized decline in FEV1.0 than bezafibrate or fenofibrate. Although the underlying mechanism remains unclear, this difference may be related to the stronger pharmacological activity of pemafibrate as a selective PPARα modulator. Together with our *in vitro* and *in vivo* experimental results, these results suggest that the senotherapeutic effects of pemafibrate through the PPARα–TFEB axis may contribute to preservation of FEV_1.0_ during aging.

This study has several limitations. First, pemafibrate was administered at the beginning of the CS exposure in our mouse model. To further clarify the efficacy of pemafibrate as the treatment for established COPD, pemafibrate should be administered after development of the COPD phenotype as well in mouse models(45). Second, while we chose oral administration as the drug delivery route in this study, the efficacy of pemafibrate upon lung local administration remains uncertain. If the effect of pemafibrate can be exerted by means of lung administration, inhalation therapy could be an effective treatment modality for respiratory diseases, since it serves as a specific local therapy without systemic adverse events; this should be examined in the future. Additionally, although pemafibrate administration in the COPD model was based on drug concentrations used in preclinical studies for hyperlipidemia and the clinical safety of pemafibrate has already been established as a treatment for hyperlipidemia at the current dose, supporting the notion of safer drug repositioning, potential adverse effect risks should be carefully monitored especially in current smoking COPD cases. Third, although we propose that the anti-senescence effects of pemafibrate on epithelial cells, mediated through the regulation of mitophagy, represent an important mechanism of action, multiple cell types—including immune, endothelial, and fibroblast populations—also contribute to COPD pathogenesis. Although our immunohistochemical analyses demonstrated increased TFEB expression in airway epithelial cells in both human and mouse lungs after pemafibrate treatment, a limitation of this study is that bulk lung RNA-seq reflects heterogeneous cell populations and does not permit precise attribution of transcriptional changes to specific cell types, including alveolar and airway epithelial, immune, or stromal compartments. Therefore, the observed changes in autophagy- and TFEB-related signatures may reflect combined effects across multiple cell types and should be interpreted with caution when inferring epithelial-specific mechanisms. Future studies using single-cell transcriptomics or cell type–specific genetic models will be required to more precisely define the relative contributions of epithelial and non-epithelial compartments. Fourth, our cohort was a retrospective analysis that evaluated the decline in naturally occurring FEV_1.0_ during aging. Accordingly, the beneficial role of pemafibrate in preventing disease progression in patients with COPD should be examined in a prospective cohort setting as well.

In conclusion, pemafibrate may exert senotherapeutic effects by regulating the TFEB–autophagy/mitophagy–lysosome axis and may represent a novel and promising therapeutic agent for COPD.

## Methods

### Sex as a biological variable

Our animal study examined female mice because female mice exhibit obvious phenotype in this experimental model. Our cohort study examined both male and female, and sex was included as covariates for statistical analysis.

### Cell culture, antibodies, and reagents

Normal and COPD lung tissues were collected from pneumonectomy and lobectomy specimens of primary lung cancer, as described previously(2). COPD was diagnosed according to the criteria of the Global Initiative for Chronic Obstructive Lung Disease(47). HBECs were isolated by means of protease digestion, cultured on rat tail collagen type I-coated (10 mg/ml) dishes in bronchial epithelial growth medium (Lonza, CC-3170). HBECs were serially passaged until passage three and then used for the experiments. Primary HBECs from non-COPD donors were used for CSE and pemafibrate experiments. The bronchial epithelial cell line BEAS-2B was cultured in RPMI 1640 medium (Thermo Fisher Scientific, 11875-093) with 10% fetal bovine serum (Cytiva, SH30910.03) and penicillin-streptomycin (Thermo Fisher Scientific, 15070-063).

The antibodies used were mouse anti-CDKN1A (NBP-29463) from Novus Biologicals; rabbit anti-CDKN1A (2947), anti-CDKN2A (29271), anti-TFEB (4240), anti-LC3B (3863), and anti-pH2A.X (2577) from Cell Signaling Technology; mouse anti-TOMM20 (sc-17764) and anti-ACTB (sc-47778) from Santa Cruz Biotechnology; and Rabbit anti-TFEB (13372-1-AP) from ProteinTech. Antibody dilutions were prepared according to the manufacturer’s instructions. The following reagents were used: pepstatin A (Peptide Institute, 4397), E64d (Peptide Institute, 4321-v), bafilomycin A1 (Sigma-Aldrich, B1793), torin1 (Selleck Chemicals, S2827), Hoechst 33258 (Fujifilm Wako, 346-07951), and DAPI (Thermo Fisher Scientific, R37606,). Pemafibrate was provided by Kowa Company, Ltd.

### Plasmids, siRNA, and transfection

LC3B complementary DNA (cDNA), a kind gift from Dr. Mizushima (Tokyo University, Tokyo, Japan) and Dr. Yoshimori (Osaka University, Osaka, Japan), was cloned into the pEGFP-C1 vector. The pEGFP-N1-TFEB vector (#38119, Shawn Ferguson Lab), the ptfLC3 vector (#21074, Tamotsu Yoshimori Lab), and the mito-dendra2 vector (#55796, David Chan Lab) for *E. coli* DH5a cells was purchased from Addgene (https://www.addgene.org/). These plasmids were transfected into BEAS-2B cells using Lipofectamine™ 3000 (Thermo Fisher Scientific, L3000015), and stably expressing clones were selected by culturing in G418 (Fujifilm Wako, 070-05183; 1.0 mg/ml)-containing medium. siRNAs targeting TFEB (s15495), PPARα (s10880), and negative control siRNAs (AM4635) were purchased from Thermo Fisher Scientific. These cells were also transfected into BEAS-2B cells or HBECs using Lipofectamine™ 3000 and Opti-MEM™ (Thermo Fisher Scientific, 31985062).

### RNA isolation and qRT-PCR

Total RNA was extracted from cultured cells or mouse lungs using QIAzol (Qiagen, 1023537) and the miRNeasy Mini Kit (Qiagen, 217004). RNA quantity and quality were evaluated using a Nanodrop™ ND-1000 spectrophotometer (Thermo Fisher Scientific) and an Agilent Bioanalyzer (Agilent Technologies), prior to bulk RNA-seq. For mRNA expression analysis using qPCR, cDNA was generated from total RNA using a Reverse Transcription Reagent Kit (Takara Bio, RR037A). Real-time PCR was performed using the SYBR™ Green™ method, as described previously(2). Data were collected and analyzed using QuantStudio™ 3 and QuantStudio™ Design & Analysis Software version 1.5.1 (Thermo Fisher Scientific). Relative mRNA expression was calculated using the 2−ΔΔCt method, with ACTB as the reference gene and the indicated control group as the calibrator.

The primers (human primers, unless otherwise noted) used were TFEB sense primer, 5’-ACCTGTCCGAGACCTATGGG-3’; TFEB antisense primer, 5’-CGTCCAGACGCATAATGTTGTC-3’; mice TFEB sense primer, 5’-AAGGTTCGGGAGTATCTGTCTG-3’; mice TFEB antisense primer, 5’-GGGTTGGAGCTGATATGTAGCA-3’; BECN1 sense primer, 5’-CCATGCAGGTGAGCTTCGT-3’; BECN1 antisense primer, 5’-GAATCTGCGAGAGACACCATC-3’; VPS18 sense primer, 5’-CACTCGGGGTATGTGAATGCC-3’; VPS18 antisense primer, 5’-TCGGAAGGGGTGAAGTCAATG-3’; ATP6V1A sense primer, 5’-GGGTGCAGCCATGTATGAG-3’; ATP6V1A antisense primer, 5’-TGCGAAGTACAGGATCTCCAA-3’; LAMP1 sense primer, 5’-TCTCAGTGAACTACGACACCA-3’; LAMP1 antisense primer, 5’-AGTGTATGTCCTCTTCCAAAAGC-3’; ATG7 sense primer, 5’-CTGCCAGCTCGCTTAACATTG-3’; ATG7 antisense primer, 5’-CTTGTTGAGGAGTACAGGGTTTT-3’; ATG10 sense primer, 5’-AGACCATCAAAGGACTGTTCTGA-3’; ATG10 antisense primer, 5’-GGGTAGATGCTCCTAGATGTGAC-3’; ACTB sense primer, 5’-CATGTACGTTGCTATCCAGGC-3’; and ACTB antisense primer, 5’-CTCCTTAATGTCACGCACGAT-3’. mice ACTB sense primer, 5’-GTGACGTTGACATCCGTAAAGA-3’; and mice ACTB antisense primer, 5’-GCCGGACTCATCGTACTCC-3’. The PCRs for TFEBs, VPS18, ATP6V1A, LAMP1, and ACTB were validated using two different primers. The primer sequences were obtained from PrimerBank (https://pga.mgh.harvard.edu/primerbank/).

### Evaluation of microarray and single-cell transcriptome datasets

We evaluated autophagy-related gene transcription in the public microarray datasets, GSE106986 and GSE994, using GEO2R (https://www.ncbi.nlm.nih.gov/geo/geo2r/). Furthermore, we analyzed recent single-cell transcriptome data sets(28). For the single-cell transcriptomic analysis, epithelial cell populations were extracted based on the cell type annotations provided in the original study.

### Preparation of CSE

CSE was prepared as described previously, with minor modifications(12). Fifty milliliters of CS were drawn into the syringe and slowly bubbled into sterile phosphate-buffered saline (PBS) in 15-ml Falcon Tubes (Thermo Fisher Scientific, 339650). One cigarette was used to prepare 10 ml of the solution. To remove insoluble particles, the CSE solution was filtered (0.22 μm; Merck Millipore, SLGS033SS); the resulting solution was designated as a 100% CSE solution.

### Fluorescence-based assessment of mitochondrial function

Mitochondrial ROS production and mitochondrial membrane potential were assessed using MitoSOX™ Red (Thermo Fisher Scientific, M36008) and MitoTracker™ Red CMXRos (Thermo Fisher Scientific, M7512), respectively, according to the manufacturers’ instructions. Fluorescence images were acquired using a fluorescence microscope (Keyence, BZ-X800), and fluorescence intensity was quantified using ImageJ. Corrected total cell fluorescence (CTCF) was calculated as integrated density − (area of selected cells × mean background fluorescence). CTCF values were normalized to the control group, which was set to 1, and expressed as relative fluorescence intensity.

### Evaluation of autophagy flux and mitochondrial turnover

BEAS-2B cells stably expressing the mRFP–GFP tandem fluorescent-tagged LC3 (tfLC3) were cultured on chamber slides and pretreated with or without 100 nM pemafibrate for 24 h, followed by exposure to 2% CSE for 1 h. Confocal imaging was performed using a laser scanning microscope (Carl Zeiss, LSM 980). To evaluate autophagic flux, the ratio of red to yellow puncta per cell was quantified using ImageJ. BEAS-2B cells stably expressing mito-Dendra2 were seeded in culture dishes and photoconverted by illumination with violet light through a DAPI filter for 6 min using a fluorescence microscope (Keyence, BZ-X800) immediately before treatment with 2% CSE for 3 h, with or without 100 nM pemafibrate. Confocal imaging was performed using a laser scanning microscope (Carl Zeiss, LSM 980). To assess mitochondrial turnover, the red fluorescence intensity was quantified using ImageJ.

### Evaluation of mitochondrial function

Mitochondrial respiration was measured using a Seahorse XF HS Mini Extracellular Flux Analyzer (Agilent Technologies) and the Seahorse XFp Cell Mito Stress Test Kit (Agilent Technologies, 103010-100). HBECs were seeded at the same density across all wells and pretreated with or without 100 nM pemafibrate for 24 h, followed by exposure to 2% CSE for 1 h before the assay. Before measurement, cell number and confluence were visually confirmed by phase-contrast microscopy, and no apparent differences were observed among the groups. Each experiment included three technical replicate wells per condition, and the experiment was independently repeated five times. For statistical analysis, values from technical replicate wells were averaged within each independent experiment, and the five independent experiments were treated as biological replicates.

### Measurement of the pH of lysosomes

For ratiometric measurement of lysosomal pH, HBECs were seeded in 35-mm glass-bottom dishes one day before live-cell imaging and pretreated with or without 100 nM pemafibrate for 24 h, followed by exposure to 2% CSE for 2 h. Lysosomal pH was assessed using LysoSensor Yellow/Blue DND-160 as previously described with minor modifications(48). Cells were then incubated with 2 μM LysoSensor Yellow/Blue DND-160 (Thermo Fisher Scientific, L7545) in complete medium for 5 min, washed twice with HBSS, and immediately imaged using a confocal microscope. LysoSensor fluorescence was detected with excitation at 405 nm and emission collected at 430–470 nm for the blue channel and 510–560 nm for the yellow channel. For pH calibration, buffers containing monensin were used to generate a standard pH calibration curve. Fluorescence intensities in the blue and yellow channels were quantified using ImageJ, and the blue-to-yellow fluorescence intensity ratio was converted to lysosomal pH values based on the calibration curve. To assess lysosomal acidity, LysoTracker™ Red staining (Thermo Fisher Scientific, L7528) was performed in HBECs grown in 12-well culture plates according to the manufacturer’s instructions, and fluorescence intensity was quantified using Image J.

### Evaluation of TFEB nuclear translocation

BEAS-2B cells expressing EGFP-TFEB were treated with 1%–5% CSE for 24 h, with or without 100 nM pemafibrate, to assess the nuclear translocation of TFEB. Torin1, a mammalian target of rapamycin inhibitor, was added to the cells at a concentration of 250 nM for 6 h before imaging. Nuclear translocation of EGFP-TFEB was quantified using ImageJ. Nuclear regions of interest (ROIs) were defined based on DAPI staining, and whole-cell ROIs were manually outlined. Cytoplasmic ROIs were generated by subtracting nuclear ROIs from whole-cell ROIs using the XOR function in the ROI Manager. After background subtraction, mean EGFP fluorescence intensity was measured in the nuclear and cytoplasmic ROIs, and nuclear translocation was expressed as the nuclear-to-cytoplasmic fluorescence intensity ratio.

### Evaluation of antioxidant pathways

Glutathione and glutathione peroxidase levels were measured in HBECs using GSH-Glo™ Glutathione Assay (Promega, V6911), and glutathione peroxidase assay kit (Abcam, ab102530), following the manufacturer’s instructions.

### Animal models

C57BL/6J mice were purchased from CLEA Japan and maintained in an animal facility at The Jikei University School of Medicine. Six to eight-week-old mice were used in all experiments. In the pemafibrate group, mice were administered 0.0003% pemafibrate mixed with normal diet, from the start of CS-exposure to sacrifice. Pemafibrate dose was determined based on the previous report and mouse blood concentration measurement data(49).

### CS exposure

Mice were exposed to a whole-body exposure system (SCIREQ, InExpose) in a barrier facility. Mice were exposed to total suspended particles of 200 mg/m^3^ using research cigarettes (University of Kentucky 3R4F research cigarettes), five days a week, for six months. Air-exposed mice served as the non-smoking controls. After six months of CS exposure, the mice underwent respiratory function tests (see below) and were sacrificed immediately. In mice that did not undergo RNA-seq analysis, bronchoalveolar lavage using 1 ml of PBS + 1% bovine serum albumin was performed before removing the lungs. The left main and right upper lobe bronchi were ligated, and the rest of the right lung was inflated at a constant pressure of 20 cm for 5 min, before fixation in 10% formalin for 24 h. Each formalin-soaked lung lobe was bisected and paraffin-embedded. Samples of the right upper lobes were snap-frozen in liquid nitrogen for RNA-seq or embedded in Optimal Cutting Temperature compound (Sakura Finetek, 4583) for histological analysis, and then stored at –80°C.

### Measurement of mouse lung function parameters

On the day of the experiment, the mice were weighed and anesthetized by means of an intraperitoneal injection of a mix of 0.3 mg/kg medetomidine hydrochloride, 4 mg/kg midazolam, and 5 mg/kg butorphanol tartrate. Once anesthesia was administered, the mice were tracheostomized using an 18-gauge metal cannula. The mice were connected to a flexiVent FX system (SCIREQ) and mechanically ventilated. Pancuronium bromide (1 mg/kg) was administered intraperitoneally, and deep lung inflation to 30 cm H2O for 3 seconds was performed before measurement. Lung function was assessed using forced oscillation and forced expiration maneuvers with FlexiWare version 8 software (SCIREQ).

### Morphometric analysis

Histological sections were stained with hematoxylin and eosin to assess alveolar enlargement. Ten random fields were evaluated by a blinded investigator using digital imaging software (ImageJ(50)), and alveolar size was estimated using the mean linear intercept method, as described previously(51).

### Next-generation sequencing and bioinformatics

cDNA libraries for RNA-seq were established from total RNA using the NEBNext^®^ Poly(**A**) mRNA Magnetic Isolation Module (New England Biolabs) to select poly-A mRNA, followed by strand-specific library preparation using MGIEasy RNA Directional Library Prep Set V2.0 (MGI Tech, 1000006386). Paired-end sequencing with a read length of 150 bases was performed on a DNBSEQ-G400 (MGI Tech) platform, following the manufacturer’s instructions. Raw sequence quality was checked using the FastQC software (http://www.bioinformatics.babraham.ac.uk/projects/fastqc/). Raw data were converted to an index file for Kallisto using the FASTA format reference transcriptome (GRCm38). Based on this index, Kallisto was run and quantified for each FASTQ dataset. Output files of Kallisto were converted to an expression matrix by the ‘tximport’ and ‘biomaRt’ packages in R. The data were normalized using the trimmed mean of M values method implemented in the edgeR package in RStudio. Following normalization, gene set enrichment analysis was performed using GSEA software v4.3.3 (https://www.gsea-msigdb.org/gsea/index.jsp).

### Western blotting

HBECs and BEAS-2B cells were lysed in M-PER Mammalian Protein Extraction Reagent (Thermo Fisher Scientific, 78510), and mouse lung tissues were homogenized in T-PER Tissue Protein Extraction Reagent (Thermo Fisher Scientific, 78501) supplemented with protease (Roche, 5892970001) and phosphatase inhibitors (Roche, 4906845001). Total protein was quantified using the bicinchoninic acid protein assay (Thermo Fisher Scientific, 23225). Western blot was performed as described previously(12). Equal amounts of protein were separated by 12.5%–15% SDS-PAGE, transferred to polyvinylidene difluoride membranes (Millipore, ISEQ00010), and incubated with the indicated primary antibodies, followed by anti-rabbit (7074) or anti-mouse (7076) IgG horseradish peroxidase-linked secondary antibody (both from Cell Signaling Technology). Proteins were detected by chemiluminescence using a ChemiDoc™ Touch Imaging System (Bio-Rad Laboratories). LC3-II was detected by means of western blot in the presence of protease inhibitors (E64d and pepstatin A), to prevent further degradation, as described previously(52).

### Immunohistochemistry and immunofluorescence staining

Immunohistochemical staining on the paraffin-embedded lung tissues was performed as described previously(22). TFEB immunostaining was assessed in terms of the ratio of positively stained cells in the small airways, and ATG5 and PINK1 immunostaining were assessed in terms of percentage of 3,3’-Diaminobenzidine (DAB) positive staining areas using ImageJ software. Immunofluorescence staining was performed as described previously, with minor modifications(22). BEAS-2B cells expressing EGFP-LC3B were grown on 8-well culture slides and treated with 2% CSE for 24 h and 20 μM bafilomycin A1 for 6 h. BEAS-2B cells were then fixed with 4% paraformaldehyde for 15 min, followed by permeabilization with 0.2% Triton™ X-100 (Fujifilm Wako, 16024751) for 10 min. After blocking with Blocking One (Nacalai Tesque, 03953-95) for 30 min, primary and secondary antibodies were applied according to the manufacturer’s instructions. Confocal laser scanning microscopy was performed using a fluorescence microscope (Carl Zeiss, LSM880). Fluorescence microscopic analysis of phosphohistone H2A.X was performed in HBECs using a fluorescence microscope (Keyence, BZ-X800, Osaka, Japan). Colocalization was quantified within cell ROIs using Pearson’s correlation coefficient with the Coloc 2 plugin in ImageJ.

### SA-β-Gal and SPiDER-β-Gal staining

SA-β-Gal staining was performed on HBECs grown in 6-well culture plates, according to the manufacturer’s instructions (Sigma-Aldrich, CS0030). Cryosections of the mice lungs (∼6 μm) were fixed in 4% paraformaldehyde for 20 min at room temperature, washed with PBS, and immersed in 20 μM SPiDER-β-Gal staining solution (Dojindo, SG02) for 1 h, at 37°C. The cells were then washed with PBS and imaged.

### Electron microscopy

Lung tissues from cigarette smoke-exposed mice were fixed with 2% glutaraldehyde in 0.1 M phosphate buffer (PB) (pH 7.3) overnight at 4℃ and then post fixed with 1% osmium tetroxide in the same buffer at 4℃ for 2 h. Dehydration was carried out using a graded series of ethanol, then the specimens were placed in propylene oxide, and subsequently embedded in Epok 812 (Oken). Ultrathin sections were prepared with a diamond knife, and then stained with uranium acetate and lead citrate, and observed by JEM-1400Plus (JEOL) electron microscope at 100 kV.

### Assessment of cell death and cell viability

Cell death was assessed by measuring the number of apoptotic cells using the TUNEL assay, which was performed using the DeadEnd™ Fluorometric TUNEL System (Promega, G3250). The TUNEL-positive cells in the lungs were detected using fluorescence microscopy (Keyence, BZ-X800). The average number of dead cells was assessed by means of manual counting of TUNEL-positive cells.

### Clinical retrospective study and data collection

We conducted a retrospective cohort study using the Standardized Structured Medical Information eXchange database(53) and electronic medical records from Jikei University Hospital, Jikei University Daisan Hospital, Jikei University Kashiwa Hospital, and Jikei University Katsushika Medical Center. Patients prescribed pemafibrate, bezafibrate, or fenofibrate between June 1, 2018, and January 31, 2023, were screened. Eligible patients had at least 2 spirometry measurements obtained under clinically stable conditions during fibrate treatment. The interval between measurements was required to overlap with treatment with the index fibrate for at least half of the observation period and not to overlap with treatment with another fibrate. Patients who received the index fibrate for less than 4 weeks between measurements or had an annualized FEV1.0 change exceeding ±180 mL/year were excluded, based on a previous benchmark study (54). Pulmonary function was measured using a spirometer (Spiroshift™ SP-790COPD; Fukuda Denshi, Tokyo, Japan). Baseline data included age, sex, BMI, smoking status and pack-years, respiratory comorbidities (COPD and bronchial asthma), hypertension, diabetes, coronary artery disease, and the use of inhalation therapy, statins, and metformin. Comorbidities were identified from physician-based diagnoses in medical records, in combination with disease-specific medications. Coronary artery disease defined as a history of angina pectoris or myocardial infarction, which was identified from physician-based diagnoses in the medical records, in combination with disease-specific medication. Inhalation therapy includes inhaled corticosteroids, long-acting beta-agonist inhalants, long-acting muscarine antagonist inhalants, and multiple combination inhalation therapy for COPD or bronchial asthma.

Forced vital capacity, vital capacity, FEV1.0, FEV1.0/forced vital capacity, and percent predicted FEV1.0 were collected at baseline and at the last eligible spirometry assessment.

### Statistics

Experimental studies are presented as mean ± SEM from at least three independent experiments. Comparisons between two groups were performed using Student’s t test, and comparisons among multiple groups were performed using one-way ANOVA followed by Bonferroni’s post hoc test.

Clinical data are presented as numbers and percentages, means and SDs, or medians and IQRs, as appropriate. Baseline characteristics were compared among the 3 fibrate groups using 1-way ANOVA for parametric continuous variables, the Kruskal–Wallis test for nonparametric continuous variables, and the chi-square test for categorical variables. The annual spirometry change was calculated from the difference between the baseline and last spirometry data; the difference was divided by the observation years between the two spirometry dates and presented as mL/year. ANCOVA was used to compare the annualized change in FEV1.0 among the pemafibrate, bezafibrate, and fenofibrate groups. The selection of covariates to be adjusted was based on previous studies and clinical relevance(43–45) and included age, sex, BMI, smoking index, baseline FEV_1.0_, comorbidities (COPD, asthma, hypertension, diabetes, and coronary artery disease), statins and metformin use, and inhalation therapy.

Experimental analyses were performed using Prism version 9 (GraphPad), and clinical analyses were performed using Stata software version 17.0 (StataCorp). All statistical tests were 2-sided, and a P value of less than 0.05 was considered statistically significant.

### Study approval

Human lung tissues, including those used for HBEC isolation, were obtained from patients undergoing pneumonectomy or lobectomy for primary lung cancer. Informed consent was obtained from all participants who underwent surgery, as part of an approved ongoing research protocol by the Ethics Committee of The Jikei University School of Medicine (Tokyo, Japan) [approval no. 20-153(5443)]. All animal experimental protocols used in this study were approved by the Jikei University School of Medicine Animal Care Committee [approval no. 2022-061]. The retrospective cohort study was approved by the Ethics Committee of The Jikei University School of Medicine [approval no. 34-171(11322)]. Based on the ethics guidelines of The Jikei University, informed consent was waived because of the retrospective study design, and an opt-out consent statement was posted on the hospital website. All clinical data were anonymized to protect participant privacy. The study was conducted in accordance with the principles of the Declaration of Helsinki.

## Data Availability

Data availabitily
Raw and processed RNA-seq data generated in this study are being deposited in the NCBI Gene Expression Omnibus. The accession number will be provided upon revision.

## Data availabitily

Raw and processed RNA-seq data generated in this study are being deposited in the NCBI Gene Expression Omnibus. The accession number will be provided upon revision.

## Author contributions

Sachi M and Saburo I performed the in vivo experiments. Sachi M and YH performed the in vitro experiments. Sachi M collected the RNA-seq data and performed the RNA-seq. Sachi M, TK, and MY analyzed the RNA-seq data. Sachi M and Saburo I collected the clinical data. Saburo I and MH performed the statistical analyses. TO and TN provided the patient samples. Sachi M, Saburo I, JA, SF, SN, Shun I, MY, YF, Shunsuke M, HH, KN, and KK interpreted the data. Sachi M, Saburo I, MY, and JA conceived the experiments and drafted the manuscript. All authors have made substantial contributions to the manuscript and read and approved the final manuscript. The order of the co–first authors was determined based on their relative contributions to the study.

## Funding Support

This work was supported by the Project for the Ministry of Education, Culture, Sports, Science, and Technology KAKENHI Grant-in-Aid for Young Scientists 21K16123, Grant-in-Aid for Scientific Research B 22H03082, Grant-in-Aid for Scientific Research C 21K08213, The Jikei University Strategic Prioritizing Research Fund 2024, and a research grant from Kowa Company, Ltd. (Tokyo, Japan).

## Acknowledgement

We would like to appreciate Yoshitaka Seki (Jikei University Katsushika Medical Center), Naoki Takasaka (Jikei University Daisan Hospital), Kazuya Tone (Jikei University Kashiwa Hospital), Masayuki Saruta (Division of Gastroenterology and Hepatology, Department of Internal Medicine, The Jikei University School of Medicine), Michihiro Yoshimura (Division of Cardiology, Department of Internal Medicine, The Jikei University School of Medicine), and Rimei Nishimura (Division of Diabetes, Metabolism & Endocrinology, Department of Internal Medicine, The Jikei University School of Medicine) for their corporation in collecting clinical data. We would like to appreciate Yuki Takemura and Toshiaki Tachibana (Jikei University School of Medicine, Tokyo, Japan) for technical supports in the electron microscopy experiment.

## Conflicts of interest statement

JA is a recipient of support from a collaborative research fund from Kowa Company, Ltd. Kowa Company, Ltd. had no role in the design of the study; in the collection, analyses, or interpretation of data; in the writing of the abstract, or in the decision to present the results. Rest of the authors declare no conflict of interest. Pemafibrate was provided by Kowa Company, Ltd.

## Notes

### Author Declarations

The Ethics Committee of The Jikei University School of Medicine (Tokyo, Japan) gave ethical approval for the use of human lung tissues, including those used for human bronchial epithelial cell isolation, obtained from patients undergoing pneumonectomy or lobectomy for primary lung cancer [approval no. 20-153(5443)].Written informed consent was obtained from all participants who underwent surgery as part of this approved ongoing research protocol. The Ethics committee of The Jikei University School of Medicine (Tokyo, Japan) gave ethical approval for the retrospective cohort study [approval no. 34-171(113229). The requirement for informed consent was waived because of the retrospective study design, in accordance with the ethics guidelines of The Jikei University School of Medicine, and an opt-out notice was posted on the hospital website. All clinical data were anonymized to protect participant privacy.

