## Supplement_data for "Senotherapeutic role of pemafibrate through autophagy/mitophagy regulation in chronic obstructive pulmonary disease"

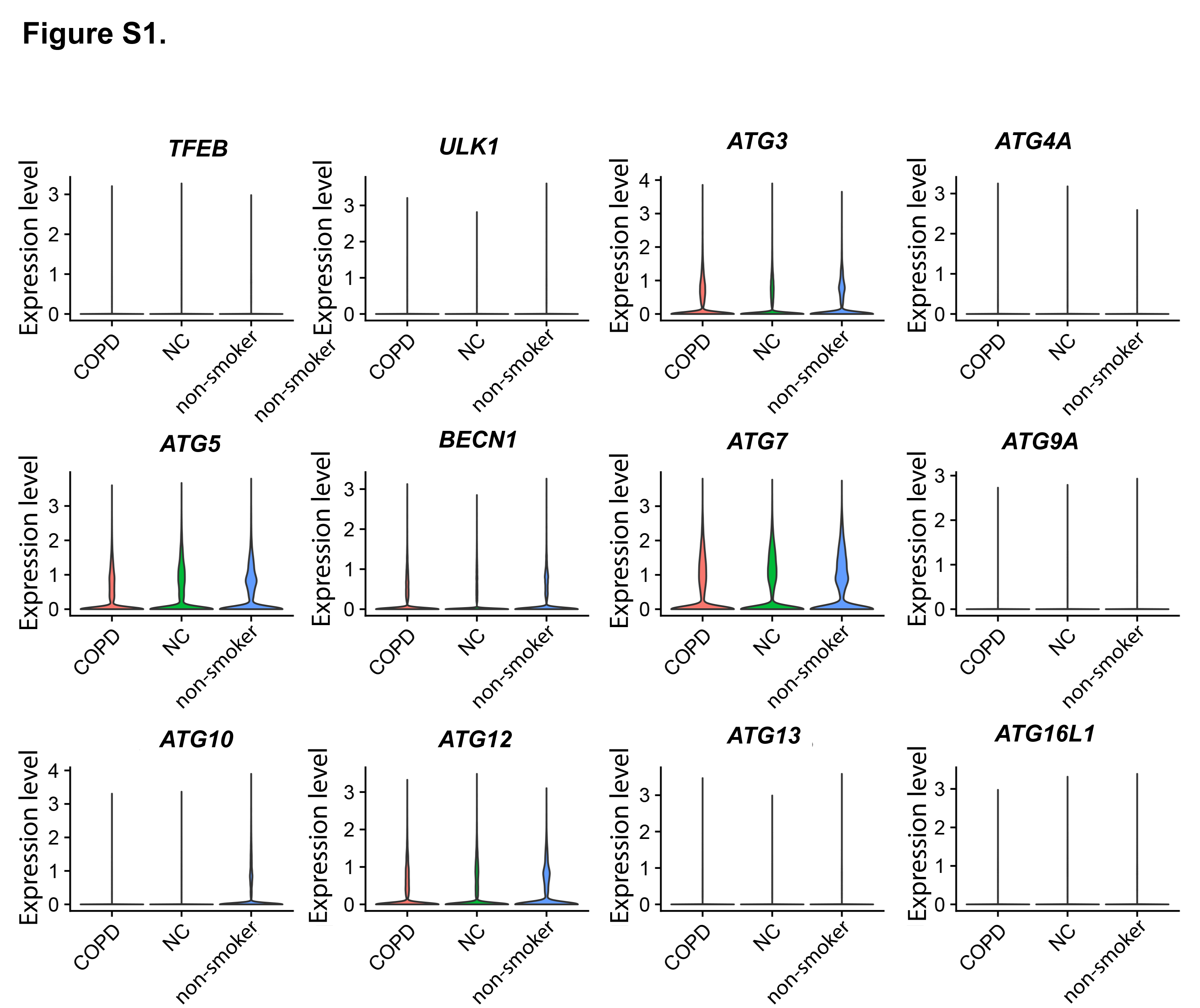

**Figure S1.** Gene expression in single-cell transcriptome datasets from all lung cells of patients with COPD, non-COPD smokers, and non-smokers.

COPD, chronic obstructive pulmonary disease; NC, non-COPD smokers

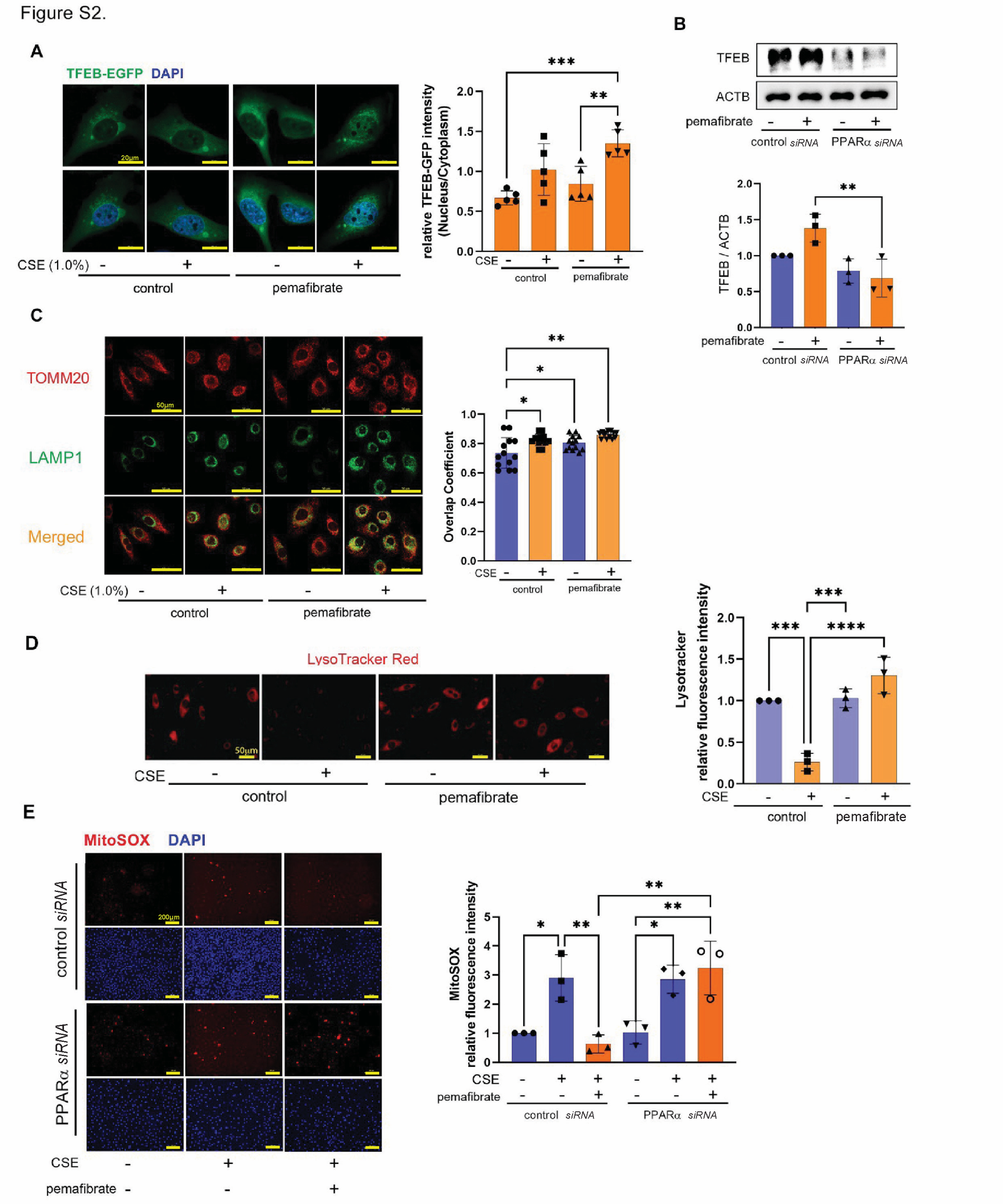
**Figure S2.**

**(A)** Fluorescence microscopy images of DAPI and EGFP-TFEB in BEAS-2B cells, after 24 h of treatment with CSE (1.0%). Pemafibrate (100 nM) was added 24 h before CSE treatment. Scale bar: 20 µm. The right panel shows the average (± SEM) relative nuclear-to-cytoplasmic EGFP-TFEB intensity ratio, quantified using DAPI-defined nuclear regions of interest (ROIs) and cytoplasmic ROIs generated from whole-cell ROIs (n = 5).

**(B)** Western blot using anti-TFEB and anti-ACTB antibodies of cell lysates of HBECs. HBECs were transfected with control or *PPARα siRNA*, and cell lysates were collected 24 h treatment with pemafibrate (100 nM). The upper panels show the representative western blot images. The lower panels show the average (± SD) of relative expression, taken from densitometric analysis (n=3).

**(C)** Colocalization of TOMM20 and LAMP1 in cigarette smoke extract (CSE)-exposed cells with or without pemafibrate treatment. Representative confocal fluorescence microscopy images showing TOMM20 (red, mitochondria), LAMP1 (green, lysosomes), and merged channels are shown for each condition. The overlap coefficient between TOMM20 and LAMP1 signals was quantified and is shown in the bar graph. Data are presented as mean ± SD. Scale bar, 50 μm.

**(D)** LysoTracker™ Red fluorescence staining of HBECs, after 24 h of treatment with CSE (2%). Pemafibrate (100 nM) was added 24 h before CSE treatment. Scale bar: 50 µm. The right panel shows the relative fluorescence intensity of lysotracker. Fluorescence intensity was quantified using corrected total cell fluorescence (CTCF). CTCF values were normalized to the control siRNA/CSE(−) group, which was set to 1, and expressed as relative fluorescence intensity (n=3).

**(E)** MitoSOX™ Red fluorescence staining of the HBECs. HBECs were first transfected with control or *PPARα siRNA*, and then 24 h later, treated with pemafibrate (100 nM), for 24 h. Finally, the cells received treatment with CSE (2%) for 24 h. Scale bar: 200 µm. The same methodology as in D was used. Data are presented as mean ± SEM from three independent experiments.

*p < 0.05, **p < 0.01, and ***p < 0.001, as analyzed using analysis of variance and Bonferroni *post-hoc* test, unless otherwise indicated.

DAPI, 4’,6-diamidino-2-phenylindole; EGFP, enhanced green fluorescent protein; TFEB, transcription factor EB; CSE, cigarette smoke extract; ACTB, β-actin; HBECs, human bronchial epithelial cells; CTCF, corrected total cell fluorescence

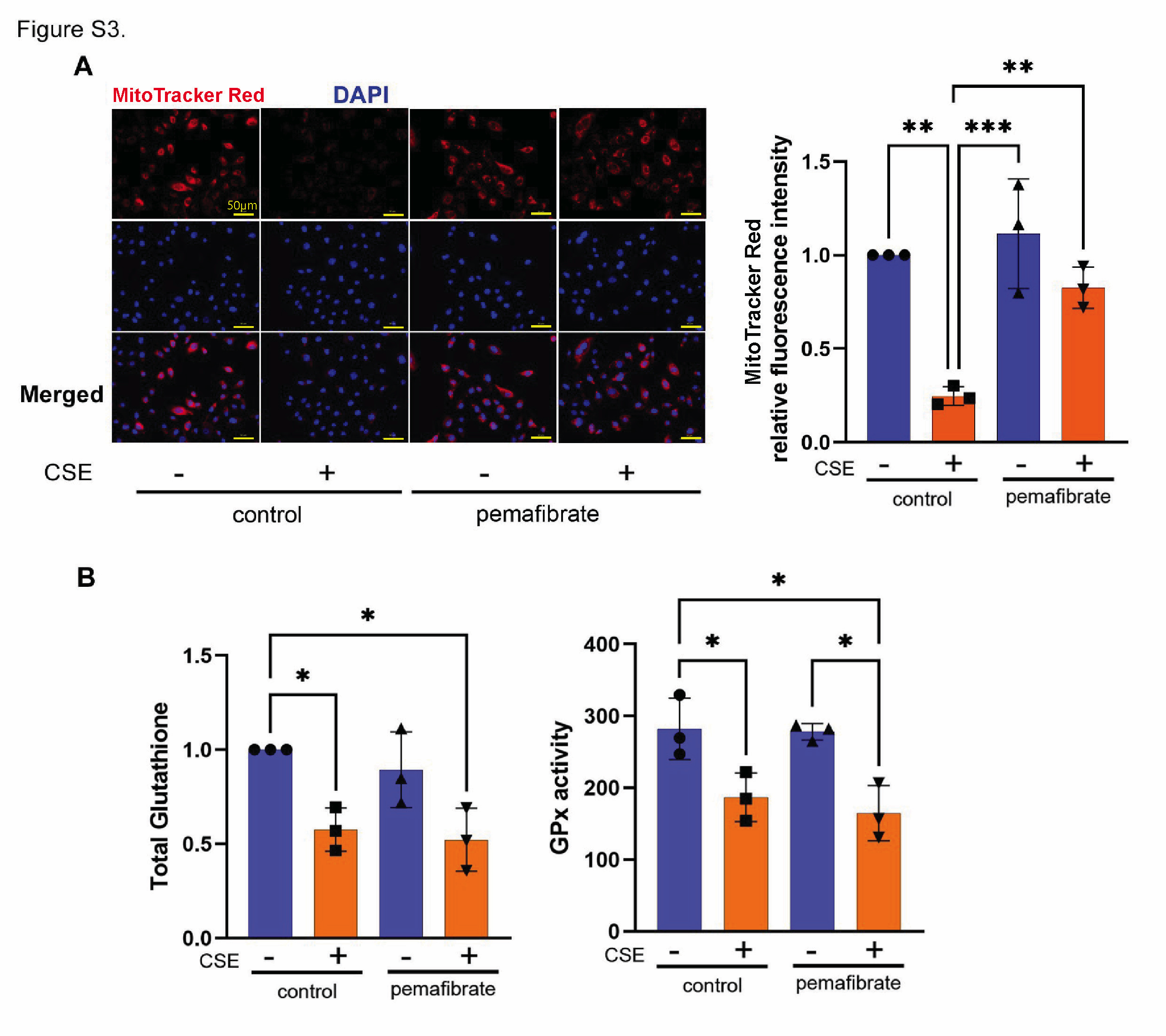

**Figure S3.**

**(A)** MitoTracker™ Red CMXRos staining of HBECs, after 24 h of treatment with or without CSE (2%) and with or without pemafibrate (100nM). Scale bar: 50 µm. The right panel shows the relative fluorescence intensity of MitoTracker Red. Fluorescence intensity was quantified using corrected total cell fluorescence (CTCF). CTCF values were normalized to the control CSE (−) group, which was set to 1, and expressed as relative fluorescence intensity (n=3).

**(B)** Evaluation of antioxidant pathway. Total glutathione (left) and glutathione peroxidase activity (right) levels of HBECs with or without CSE (2%) and with or without pemafibrate (100nM).

**p*<0.05, **p < 0.01, and ***p < 0.001, as analyzed using analysis of variance and Bonferroni *post-hoc* test.

DAPI, 4’,6-diamidino-2-phenylindole; CSE, cigarette smoke extract; HBECs, human bronchial epithelial cells; GPx, glutathione peroxidase

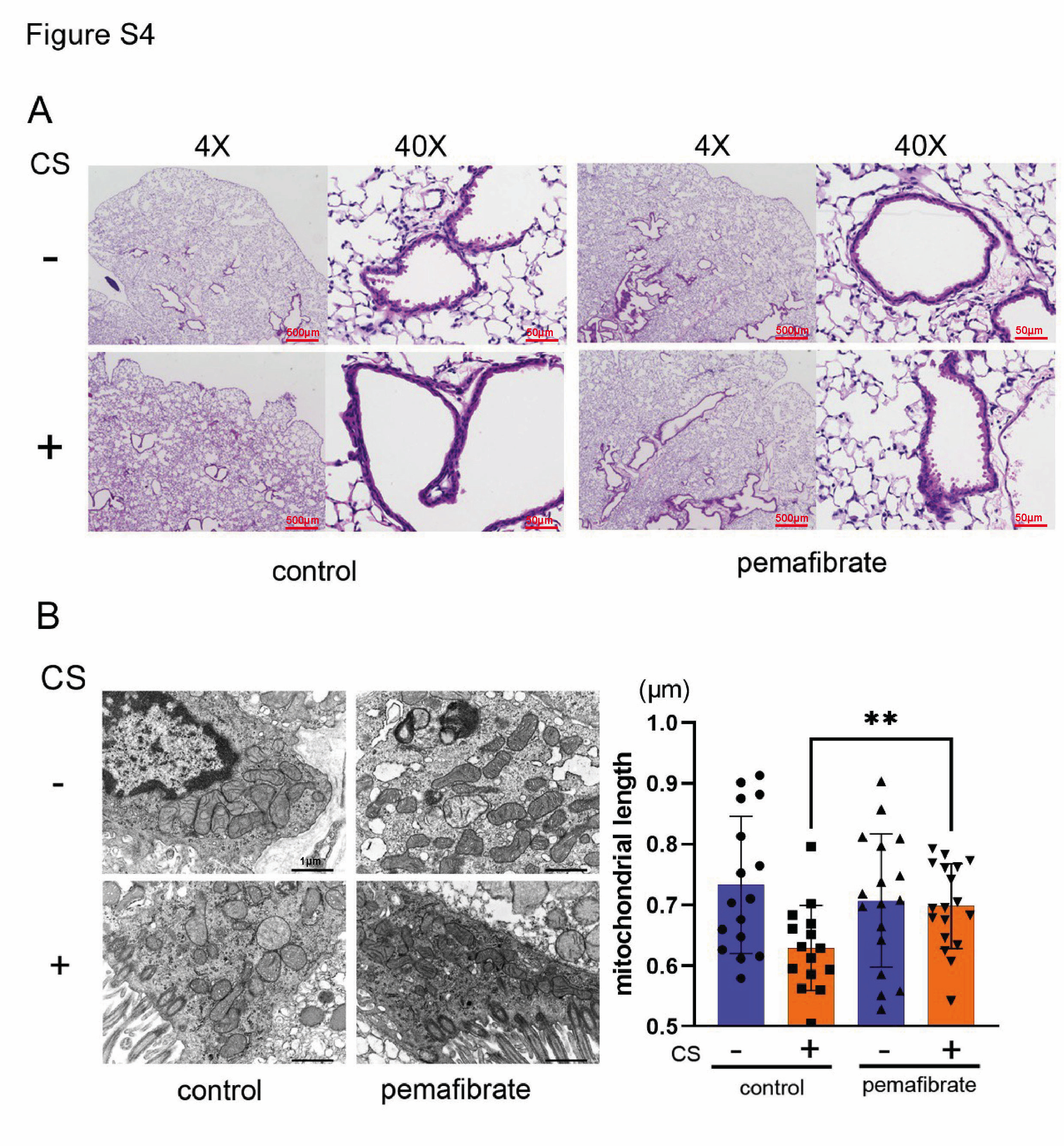

**Figure S4.**

**(A)** Histological assessment of small airway structures in cigarette smoke (CS)-exposed mice treated with pemafibrate. Representative hematoxylin and eosin (H&E)-stained lung sections from mice with or without CS exposure (CS− and CS+, respectively) treated with vehicle (control) or pemafibrate. Low-magnification (4×) images illustrate overall lung architecture, and high-magnification (40×) images highlight small airway morphology. Scale bars are indicated in each panel.

**(B)** Electron microscopy analysis of mitochondria in bronchial epithelial cells of room air or CS-exposed mice lungs. The original magnification is 12,000x, Scale Bar: 1 µm. The right panel shows the average (± SD) length of mitochondria. ***p*<0.01, as analyzed using analysis of variance and Bonferroni *post-hoc* test.

CS, cigarette smoke.

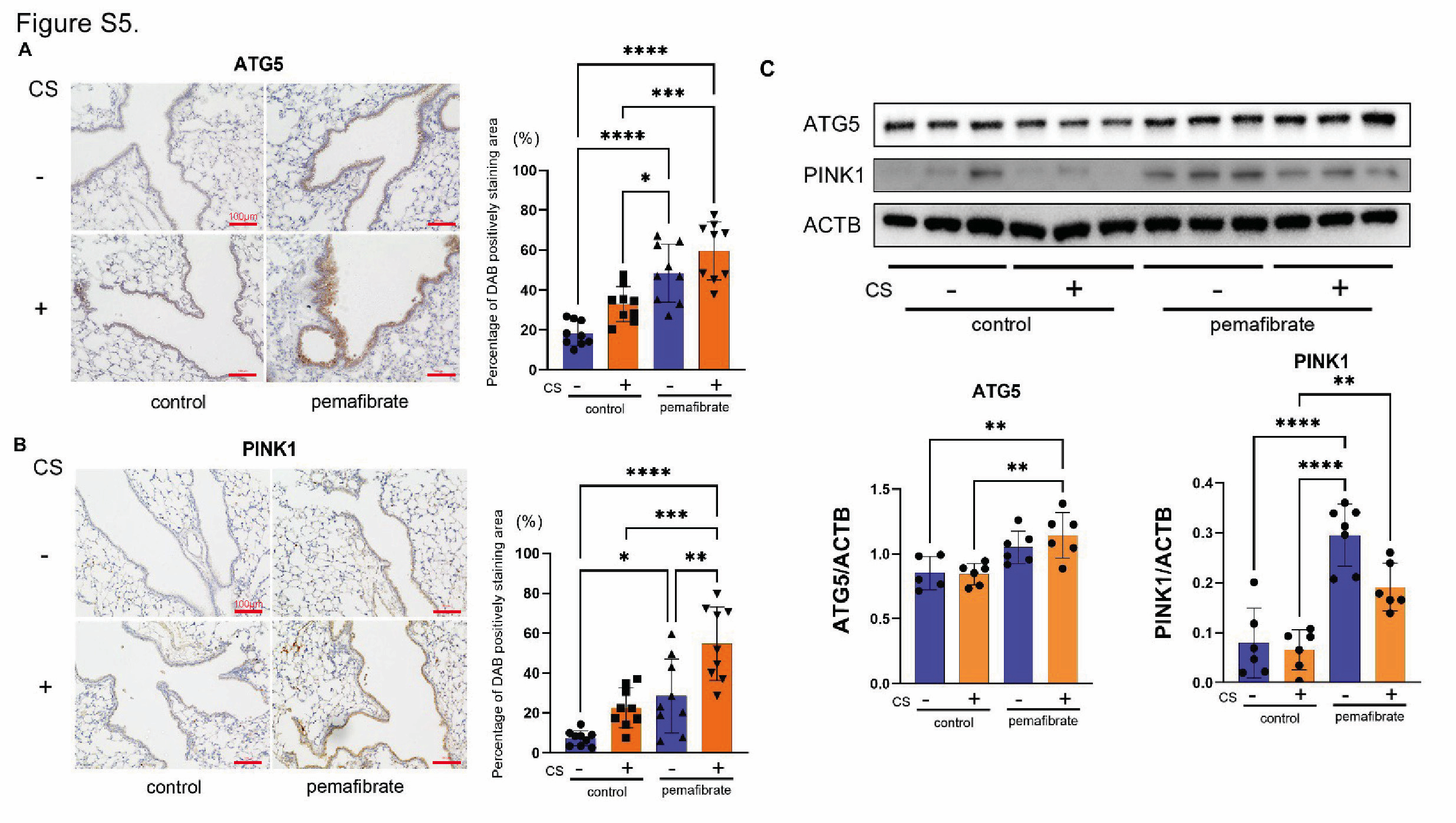

**Figure S5.**

(**A**) Immunohistochemical staining for ATG5 in the mice lung airways. Scale bar: 100 µm. The right panel shows the ratio of ATG5-positive area to the total bronchial epithelial cell area. 3 random bronchial sections were chosen blindly, and 3 mice per group were analyzed.

(**B**) Immunohistochemical staining for PINK1 in the lung airways of mice. Scale bar: 100 µm. The same methodology as in (A) was used.

**(c)** Western blots showing ATG5, PINK1, and ACTB protein levels in mouse lung homogenates. The lower panels show the average (± SD) of relative expression, taken from densitometric analysis (ATG5: n = 5-6 per group; PINK1: n = 6-7 per group).

**p*<0.05, ***p*<0.01, ****p*<0.001, and *****p*<0.0001, as analyzed using analysis of variance and Bonferroni *post-hoc* test.

ATG5, autophagy-related protein 5; PINK1, PTEN induced putative kinase 1; DAB, 3,3’-Diaminobenzidine; BEC, bronchial epithelial cells

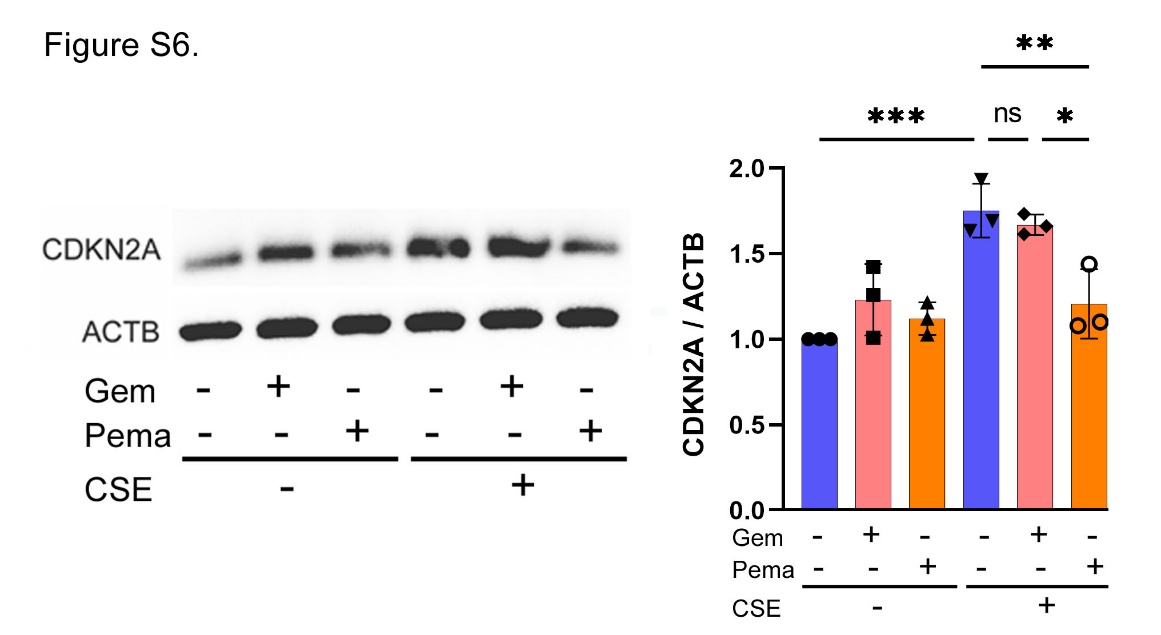

**Figure S6.** Western blot using anti-CDKN2A (p21) and anti-ACTB antibodies of cell lysates of HBECs.

Cell lysates were collected after 24 h treatment with DMSO, gemfibrozil (10μM) or pemafibrate (100 nM), with or without CSE. The left panels show the representative western blot images. The right panel shows the average (± SD) of relative expression, taken from densitometric analysis (*n*=3).

**p*<0.05, ***p*<0.01, and ****p*<0.001, as analyzed using analysis of variance and Bonferroni *post-hoc* test.

CDKN2A, cyclin-dependent kinase inhibitor 2A; ACTB, β-actin; CSE, cigarette smoke extract; Gem, gemfibrozil; Pema, pemafibrate.

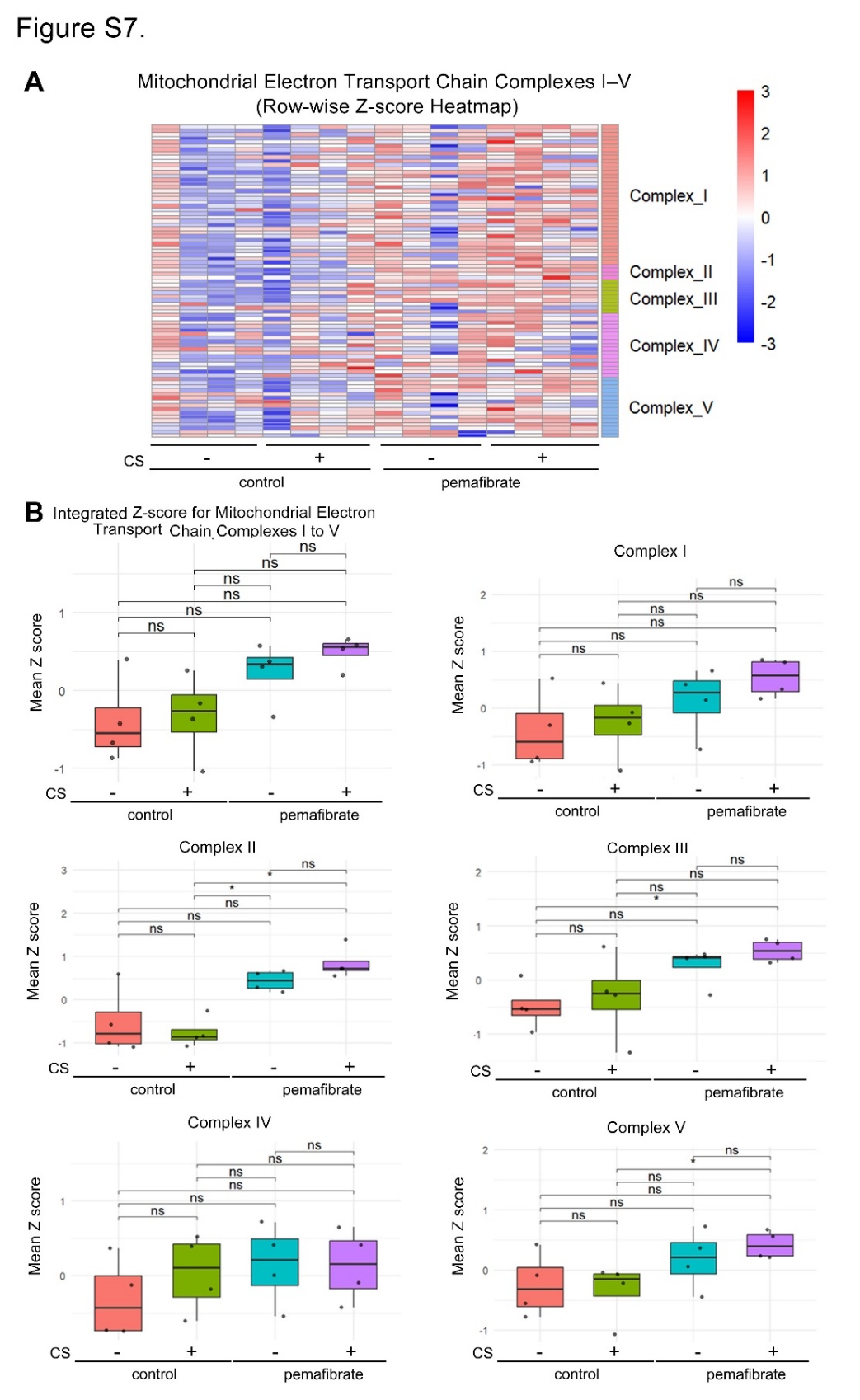

**Figure S7.** Effects of pemafibrate on mitochondrial electron transport chain gene expression.

(A) Heatmap showing row-wise Z-scores of genes involved in mitochondrial electron transport chain complexes I–V in control and pemafibrate-treated groups, with or without cigarette smoke (CS) exposure.

(B) Box plots showing integrated and complex-specific mean Z-scores (Complexes I–V). Statistical significance was analyzed using analysis of variance and Bonferroni *post hoc* test.; ns, not significant.

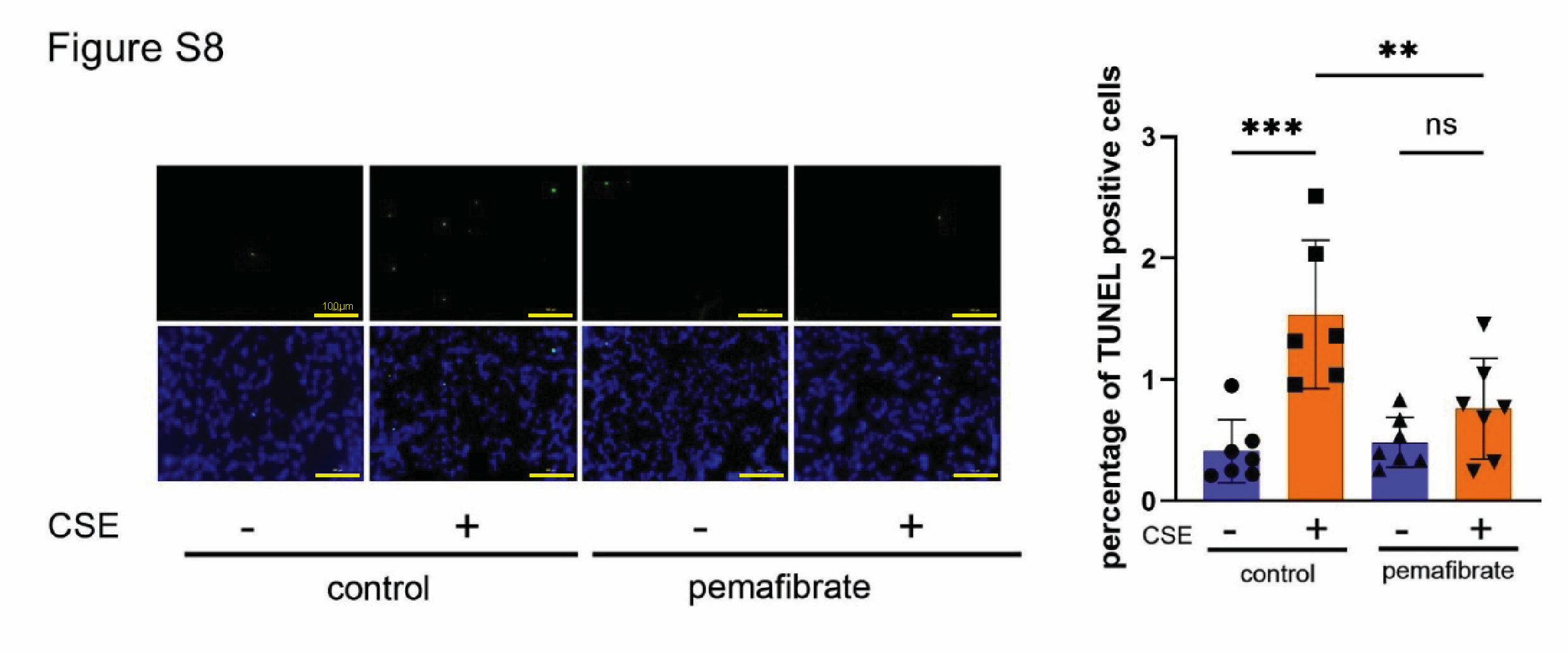

**Figure S8.** TUNEL assay staining (green) in cigarette exposed mice with pemafibrate. *n*=7 in control diet:room air, *n*=6 in control diet:CS exposure, *n*=7 in pemafibrate-mixed diet:room air, and *n*=7 in pemafibrate-miexed diet:CS exposure. The right panel show the average (± SD) of percentage of TUNEL positive cells. **p<0.01, and ***p<0.001, as analyzed using analysis of variance and Bonferroni *post-hoc* test. Scale bar: 100 µm.

TUNEL, terminal deoxynucleotidyl transferase dUTP nick end labeling; CS, cigarette smoke
